# Colorectal cancer and microbiota modulation for clinical use. A systematic review

**DOI:** 10.1101/2021.09.01.21262956

**Authors:** Julio Madrigal-Matute, Sara Bañón Escandell

**Affiliations:** Biomedical Institute for Nutrition and Health (IBIONS)

**Keywords:** cancer, colorectal, nutrition, diet, probiotic, prebiotic, symbiotic, immunotherapy, immune system

## Abstract

Colorectal cancer is one of the top contributors to the global burden of cancer incidence and mortality with genetic and environmental factors contributing to its etiology. Modifiable or environmental factors can be the cause of up to 60% of the risk of developing colorectal cancer. Hence, there is a growing interest in specifically defining what can be improved in our lifestyle to reduce this risk, improve the effectiveness of treatments, reduce side effects, and decrease the risk of recurrence. One of the elements directly related to lifestyle is gut microbiota. The microbial ecosystem has a vital role in colorectal cancer prevention and antitumoral response through modulation of the immune system and production of short-chain fatty acids. Numerous approaches have been used to identify healthy microbiota that can reduce the risk of cancer development, improve treatment efficacy, and reduce side effects.

Scientific literature in this subject is growing exponentially and, therefore, systematic reviews and meta-analysis are required to ensure that appropriate recommendations are given to patients.

This work aimed to perform a systematic analysis of the published literature to elucidate whether microbiota modulation through pre-, pro-, symbiotic treatment and/or nutritional intervention can be beneficial for patients diagnosed with colorectal cancer.

Detailed analysis of published studies shows that some prebiotics, such as inulin and resistant starch, probiotics such as lactic strains producers of short-chain fatty acids, and consumption of unprocessed plant products, can be effective recommendations for patients diagnosed with colorectal cancer. This advice should always be individually tailored and followed up by a healthcare professional with expertise in the field.

## 1. Introduction

### 1.1. Colorectal cancer

Colorectal cancer (CRC) is a disease based on an important genetic component, coupled with key influence from our lifestyle habits. Approximately 10-20% of diagnoses of CRC have a family history with the disease,^1^ and through family and twin studies, heritability is estimated to range from 12-35%.^2, 3^ Although numerous genome-wide association studies have been performed identifying genes that increase susceptibility to develop CRC,^4^ many risk factors remain unknown. However, only 5-7% of patients’ colon cancer has a clear genetic origin.^5^ Hereditary CRC syndromes can be subdivided into those associated with polyposis and those without polyposis (Lynch syndrome and familial CRC).^6^ Risk factors also include the development of inflammatory bowel disease, including Chron’s disease and ulcerative colitis. These pathologies significantly increase the chances of developing CRC, being highly dependent on environmental and nutritional factors.^7^ Specifically, geographical differences in incidence of CRC are influenced by genetic, and mainly environmental and nutritional risks.^8^

It is not surprising that environmental factors have such a strong influence on the risk of developing colon cancer, as the intestinal epithelium is one of the tissues with the highest turnover rate in our body and is in continuous direct contact with nutrients, food toxins, and microbiota.^9–14^

#### Etiology

Among the metabolic changes that precede the development of cancer are chronic cell damage due to a pathological increase in reactive oxygen and nitrogen species (ROS and RNS), DNA damage, and chronic inflammation.^6^ Inflammation triggers the release of pro-inflammatory cytokines into the gut to regenerate damaged tissue. In certain situations, inflammation becomes chronic, forming a positive feedback loop that facilitates tumor development.^15^ Colon tumors generally begin with the transformation of stem and epithelial cells that reside at the base of the colonic crypts. These cells undergo chronic damage by inflammation, oxidative stress, and/or genetic mutations that evolve into a premalignant lesion or polyp.^16^ These polyps follow two main routes of differentiation to malignancy: the *adenoma-carcinoma* type (70-90%) and the *serrated* (10-20%) type. In addition, another route not associated with polyposis has been described with a lower incidence, characterized by *microsatellite instability* (MSI; 2-7%).^17^ The development of a malignant tumor is estimated to take 10-15 years on average.^6^

### 1.2. Gut microbiota

The microbiome includes all types of microorganisms associated with the host, interacting in a complex way and usually found in the epithelial barrier zones.^18^ It is acquired at birth by vertical transmission and is modulated throughout our lives by environmental factors.^19^ The microbiota is a dynamic niche that is involved in the maintenance of health and in the origin and development of numerous diseases by modulating the homeostasis of the immune system.^20^ Although the mechanisms may be varied, one of the most notably is the stimulation of the T-cell response, through the expression of bacterial epitopes or their cross-reactivity with tumor epitopes. Additionally, gut microbiota also modulates the response to tumors by inhibiting specific receptors that promote inflammatory and immune responses. Finally, another important mechanism carried out by the microbiota is the synthesis of metabolites, such as short-chain fatty acids (SCFA), which elicit a local and systemic response in the host organism.^19^

But what is the ideal microbiota composition for health? This is a question that is not easy to answer, although numerous studies have attempted.^21, 22^ Analyzing a metabolite panel comparing stool from healthy versus CRC patients, it was found that the best marker to discern between a healthy patient and a CRC patient is primarily a decrease in the CRC patient’s levels of certain SCFA, in particular, acetate and butyrate.^23^ The protective effect of SCFA against CRC may be due to their ability to decrease inflammation and modulate the immune response.^24^ In addition, SCFA inhibit proliferation and promote apoptosis in neoplastic colonocytes.^25^

SCFA play a key role in maintaining intestinal health and preventing the development of numerous pathologies.^26^ They are produced by many bacteria in the intestinal tract, in particular, lactic acid bacteria and bifidobacteria. SCFA transporters and their receptors are the communication bridges between the microbiota and the cells of the gut.^27^ Butyrate has been shown to decrease intestinal inflammation by regulating epithelial occlusive junctions,^28^ which are essential for maintaining the integrity of the intestinal barrier and mucosa by aiding in the homeostasis of ions, nutrients and water.^29^

Among the SCFA-producing bacteria, and specifically butyrate, are those of the genus *Butyricicoccus*, which could protect against colitis in patients with irritable bowel syndrome, as these bacteria are reduced in affected patients.^30^ Likewise, other bacteria belonging to the phylum *Firmicutes* such as *Eubacterium rectale*, *Ruminococcus albus*, *R. callidus, R. bromii* and *F. prausnitzii* are 5 to 10 times more abundant in healthy patients than in patients with Crohn’s disease.^31^ The decrease in species such as *Bacteroides fragilis* and *B. vulgatus* is associated with reduced levels of butyric and propionic acid and increased inflammatory markers.^32^ Additionally, *Akkermansia muciniphila,* one of the most abundant components of the intestinal microbiota, is found to be decreased in obese patients,^33, 34^ those with ulcerative colitis,^35^ and those with CRC.^36^ *A. muciniphila* ferments mainly mucins, as well as other mono- and polysaccharides, giving rise to various SCFA.^37^

Taken together, it appears that SCFA-producing bacteria, such as acetate and butyrate are often associated with healthier microbiota profiles. For these reasons, SCFA are being tested as a therapy for CRC.^38^

#### Dysbiosis

Pathological changes in the composition of the microbiota is known as dysbiosis.^39^ This unbalance in the homeostatic microbiome is related to the origin of various types of tumors due, in part, to the modulation of tumor immunity,^40^ with CRC being prominent among these tumor types.^41^ Patients with CRC are often characterized by a decrease in beneficial microbiota and an increase in pathogenic opportunistic microorganisms.^22^ This pathogenic imbalance can have a variety of origins. A frequent and often underestimated cause of dysbiosis stems from intensive antibiotic use.^42^

Chronic intestinal inflammation and epithelial disruption, two of the most important risk factors for CRC development, are related to intestinal dysbiosis.^21, 43^ Many of the pathogenic species identified in different studies associated with the development of CRC belong to the group of hydrogen sulfide (H_2_S)-producing bacteria. These bacteria can oxidize SCFA, decreasing its levels locally.^44^ H_2_S is highly toxic to colonocytes, causes damage to the intestinal epithelium, promotes inflammation, produces metabolic changes, especially through oxidation of butyrate, and can result in the development of CRC.^45, 46^ H_2_S is also rapidly absorbed by the mucosa,^47^ which can disrupt the balance between proliferation and apoptosis^48, 49^ and lead to DNA damage.^50^ Furthermore, feces of people at high risk of CRC recurrence have higher concentrations of H_2_S.^51^ Other studies corroborate these results by finding an increase in *Fusobacterium spp.*, and more specifically *Fusobacterium nucleatum*, an H_2_S-producing species, in colonic tumor tissue.^52, 53^ *F. nucleatum* also promotes tumor growth, cell proliferation and the acquisition of chemoresistance in CRC, favoring inflammation and autophagy as a cytoprotective mechanism of the tumor.^54, 55^

At the phylum level, CRC patients show increased abundance of *Proteobacteria* and *Verrucomicrobia*.^56^ Species associated with CRC development include *Bacteroides fragilis, Escherichia coli, Peptostreptococcus spp, Enterococcus faecalis, Streptococcus bovis,* or *Porphyromonas spp.*^22, 53, 57–59^ For example, *B. fragilis* and *E. coli play* a key role in the development of familial CRC. In patients with adenomatous polyposis, areas arise in the colonic mucosa where *B. fragilis* and *E. coli* accumulate, leading to a local increase in oncotoxins such as *B. fragilis* toxin and colibactin. ^60^ These oncotoxins, when transferred to the colon of mice, increase colonic tumor growth and mortality.^60^ In addition, *B. fragilis* levels are elevated in patients with diarrhea, with chronic diarrhea being an important risk factor for the development of CRC.^61^

Nonetheless, it may not be so simple to discern which bacteria are beneficial. In general, it appears that it may be context-dependent and that one of the key characteristics of a healthy microbiota is its beneficial/harmful microbiota ratio and/or its diversity.

### 1.3. Food and gut microbiota

Due to the importance of the microbiota in health and in the origin of diseases such as CRC, there is much interest in knowing how to modify the flora in order to have a better homeostatic capacity. The main way in which we can interact directly with our microbiota is through the food we eat.^62^ A study of monozygotic and dizygotic twins, and their mothers, shows that while there is a common core microbiota, pathological conditions, such as malnutrition leading to obesity, produce a significant change in the microbiota. These changes result in reduced diversity and modifications in gene expression and metabolic pathways.^62^ Along these lines, it has been shown that a “Western” diet (high fat and sugar) can alter the microbiota in periods as short as 1-3 days, and that these changes can be reversed by following a healthier diet (high plant).^63, 64^ These results show that a healthy diet, mainly a plant-based, can improve our microbiota faster than probably any other therapeutic intervention. In fact, adherence to a Mediterranean diet has been shown to have a beneficial outcome in modulating the microbiota. Participants following this type of diet had higher levels of SCFA, *Prevotella* and *Firmicutes* that degrade plant fibers, in addition to a higher proportion of bifidobacteria/E*. coli* and better gastrointestinal habits.^65^ Therefore, it is likely that part of the health benefits of this dietary pattern are due to its effect on our commensal flora.

In fact, the health benefits of certain dietary patterns, such as fiber consumption, have been evaluated for decades. However, the mechanisms by which fiber decreases the incidence of CRC are not fully understood. Two mechanisms that may be complementary are generally postulated: i) fiber may facilitate gastrointestinal transit decreasing the contact between procancerous substances and ii) the intestinal epithelium, or gut bacteria may ferment fiber, producing SCFA.^66^ In an attempt to dissect these mechanisms, a study was conducted using a mouse model in which a representative human microbiota (gnobiotic) was “seeded”. It was found that a low-fiber diet leads to an overgrowth of bacterial species that cause degradation of the mucus lining the intestinal barrier. This makes the barrier more permeable to pathogenic organisms, facilitating the development of colitis.^67^ In this mouse model, with a low-fiber diet, overgrowth of *B. caccae* and *A. muciniphila* was promoted, with these two species being responsible for the degradation of intestinal mucus.

The importance of nutrition is often underestimated in most scientific and medical fields; however, available data show that it is a key tool in modifying the microbiota.

### 1.4. Probiotics

Probiotics are defined as “live microorganisms that confer a health benefit when administered in adequate amounts”.^68^ The benefits are varied and range from the maintenance of intestinal homeostasis to the prevention of diarrhea, irritable bowel syndrome, and constipation.^69^ In addition, numerous *in vitro*,^70^ preclinical^71–73^ and clinical studies^74^ have shown that probiotics may have beneficial effects, specifically in cases of CRC.

Probiotics exert their beneficial action by modifying the gut microbiota, improving the gut barrier and its microenvironment, secreting anti-cancer metabolites, and reducing inflammation.^75^ In addition, probiotics have an intimate relationship with the immune system, particularly through homeostasis of epithelial cell junctions and modulation of T-lymphocytes and dendritic cells activation.^36, 76^ As an example, *Butyricicoccus pullicaecorum* reduces necrotic enteritis and pathogen abundance in the caecum and ileum in broiler chickens.^77^ Furthermore, its administration promotes the proper functioning of the intestinal epithelial barrier and reduces colitis in preclinical models.^30^ One of the most widely used probiotics is *A. muciniphila*, which has been shown in numerous studies to have a beneficial role in health maintenance and a therapeutic effect on various diseases such as obesity,^78^ or epithelial tumors.^79, 80^ In this regard, it has been used in preclinical models of chronic colitis, where it improved inflammatory parameters and damage in the colon ^81^ Additionally, lactic. acid bacteria have been frequently used for their health benefits and their immunomodulatory effect,^82^ contributing to inhibiting the development of pathogens.^89^ Treatment with *L. salivary* and *L. fermemtum* in combination with *L. acidophilus* can reduce CRC cell proliferation in experimental models.^83, 84^ Furthermore, oral administration of 21 different strains of *Lachnospiraceae* attenuated colorectal inflammation and improved the composition of the microbiota in preclinical models.^85^

However, a meta-analysis evaluating the role of probiotics in cancer patients failed to find a clear overall beneficial outcome, except in reducing the degree of diarrhea, bowel movement, and the need for antidiarrheal drugs.^86^

### 1.5. Prebiotics

Prebiotics are defined as “any substance that is selectively utilized by the recipient organism and confers a health benefit”.^87^ One of the most studied prebiotics is inulin. Inulin is part of a group of dietary fibers known as inulin-type fructans. These compounds are found in numerous foods such as garlic, artichoke, onion and asparagus.^88^ Inulin-type fructans resist digestion in the small intestine and are fermented in the colon, where they are transformed into lactic acid and SCFA, improving intestinal health.^89, 90^ Inulin has been shown to increase levels of CRC-protective bacteria such as *Bifidobacteria*,^91^ *Bacteroides,*^92^ and *Akkermansia muciniphila*^78^ in murine models.

The role of resistant starch as a prebiotic has also been examined in numerous pathologies and its benefits have also been suggested in the field of cancer, particularly in CRC.^93^ Resistant starch is a group of carbohydrates that resist digestion by pancreatic amylase in the small intestine. This type of starch then reaches the colon where it is fermented by the microbiota, producing numerous protective effects against tumor development, including production of SCFA (butyrate), regulation of bowel movements and the reduction of colonic pH.^93, 94^ Many foods contain resistant starch, such as cereals, vegetables, legumes, seeds, and some nuts.^93, 95^

Recent studies are using prebiotics as a basis for the synthesis of therapeutic targets for cancer treatment because of their promising role in this field. ^96^

### 1.6. Symbiotics

Symbiotics are defined as “a mixture including live microorganisms and substrates, which used selectively by the host organism confers a health benefit”.^97^ A distinction is made between *cooperative* symbiotics, in which probiotic and prebiotic are administered together and act independently contributing to the health benefit, and *synergistic* symbiotics, where the probiotic uses the co-administered prebiotic as a substrate.^97^ The use of symbiotics is becoming increasingly common in the biomedical literature as a method of enhancing the effects of pro- and prebiotics used individually. For example, the combination of *Bifidobacterium animalis subsp. Lactis* with a resistant starch led to a decrease in the number and incidence of colon neoplasms. Interestingly, the probiotic alone did not produce a similar effect. It was the combination that resulted in protection against cancer development, in part through increased production of SCFA.^98^

Therefore, it seems very promising that symbiotics may play a more effective role than pre- or probiotics alone.

### 1.7. Oncological treatments and microbiota in CRC

Technological advances and new therapies have dramatically increased the life expectancy of CRC patients.^6^ However, CRC patients often show particularly low responses to hospital treatments.^99^ For this reason, numerous lines of research are being devoted to better understanding the mechanisms underlying this lack of response to treatments, such as immunotherapy.

The first evidence showing the key role that the microbiota plays in modulating the body’s immune response to cancer treatment came from a pioneering study showing that microbial flora, and in particular lipopolysaccharide (LPS) from Gram-negative bacteria, increased the activation of dendritic cells and TCD8+ lymphocytes. In this way, the microbiota promoted melanoma regression after radiotherapy treatment. In contrast, the use of antibiotics, inhibition of LPS and its signaling, significantly diminished the effect of radiotherapy treatment.^100^ Numerous studies have corroborated these data, showing that therapies decrease their efficacy in axenic (without microbial flora) or antibiotic-treated mice.^80, 101–103^ A key study in this field showed that genetically identical mice from two different suppliers responded unequally to the same cancer treatment, due to differences in microbiota. Not only that, but cohousing or fecal transfer eliminated these differences, clearly pointing towards a key role of microbiota in modulating the efficacy of cancer treatment.^104^

Although the role of the microbiota has been studied in different cancer treatments such as radiotherapy,^100^ chemotherapy,^102^ and surgery,^105^ most studies have focused in immunotherapy.^80, 103^ This may be due to both the microbiota’s promising role in cancer treatment and its key role in modulating the immune response.

#### Immunotherapy

Unfortunately, a large proportion of CRC patients do not respond well to immunotherapy. It is known that the effectiveness of some treatments depends, to a large extent, on the individual’s genetics and/or cancer phenotype^99^; however, for a majority of CRC patients, the cause of their lack of response is unknown.

Immunotherapy has promising results in numerous tumor types by inducing immunity by T lymphocytes ^106^ Thus, the anti-PD-1 drugs pembrolizumab and nivolumab are effective in metastatic patients whose tumors are deficient in DNA mismatch repair (dMMR) enzymes and have high microsatellite instability (MSI-H). ^99^For the treatment of refractory dMMR-MSI-H tumors, pembrolizumab and nivolumab combined with an anti-CTLA4, ipilimumab, are used. However, in tumors with active DNA repair enzymes and low microsatellite instability, which account for 85% all tumors, these therapies tend to be ineffective.^99^ Therefore, there is great interest in finding methods to improve the efficacy of these therapies in CRC.

Knowing the ideal composition of microbiota to improve the efficacy of immunotherapy treatments is no easy task. It is likely, as mentioned above, that the key lies in increasing the ratio of bacteria that are beneficial to health and decreasing those that are harmful.^107^ In this line, mice transplanted with microbiota from CRC patients show, after treatment with anti-PD-1, a decrease in the anti-tumor response in CRC and lower survival, compared to those transplanted with microbiota from healthy controls.^117^ Among the bacteria that may be beneficial to improve immunotherapy are: *Clostridiales*, *Ruminococcacae*, *Enterococci*, *Collinsella*, *Alistipes*, *Faecalibacterium spp*, *A. muciniphila*, *B. fragilis*, *Eubacterium linosum* or *Bifidobacteria*, although most studies have been performed on epithelial type tumors.^19, 79, 80^

Therefore, the study of the microbiota in the treatment of cancer patients, especially in CRC patients whose treatments do not reach the effectiveness of other types of cancer, is essential.

#### Effect of surgery on the microbiota

Colorectal surgery in humans aims for total tumor removal plus the addition of a safety perimeter. When this clean-border surgery is achieved, the likelihood of tumor recurrence decreases considerably.^108^ However, local recurrences are common, up to 15%, the origin of which appear to be linked to the biology of the primary tumor and/or seeding of the tumor bed during surgery.^109–111^ Most tumors recur at the site of the anastomosis due to incomplete wound closure, the inherent porosity of the junctional area, improper healing, and/or the presence of collagenolytic organisms that increase porosity.^112, 113^ In this regard, a high-fat diet increases the production of colorectal tumors in preclinical models of anastomosis.^105^ This mechanism is linked to the overproduction of collagenolytic organisms at the suture site. Pre-operative fecal analysis shows that obese patients respond by increasing collagenolytic micro-organisms after taking antibiotics. In contrast, non-obese patients have decreased levels of collagenolytic micro-organisms.^105^

Therefore, a healthy microbiota low in collagenolytic microorganisms is key to increasing the chances of successful CRC surgery and avoiding recurrences.

### 1.8. Study aims

The overall aim of this work is to review published studies on the modification of the microbiota in CRC.

The specific objectives of this work are to evaluate the impact of modulating the microbiota in CRC patients through; 1) probiotics, 2) prebiotics, 3) symbiotics and 4) nutritional changes. We aim to analyze the potential use of these approaches to modulate the microbiota in CRC patients.

## 2. Methods

### 2.1. Systematic search

The search strategy was defined between January 15^th^ and April 15^th^, 2021. The search and filtering in the Pubmed database took place on April 24^th^, 2021. The language used for the search and filtering of the publications was English. The detailed analysis of the selected publications was carried out from April 25^th^ to May 31^st^, 2021.

The search strategy included six types of terms: (1) target disease of the study; (2) type of cancer; (3) microbiota and dysbiosis; (4) oncology treatment, (5) use of nutrition and/or supplementation with pro-, pre- and symbiotics and (6) original articles.

To broaden the search terms, asterisks were added to the end of the words. Keywords were combined using the Boolean operators “AND”, “OR” and “NOT”. The search strategy is detailed in Table 1.

**Table 1.** Details of the search strategy used.

| Table 1. Details of the search strategy used. |  |  |
| --- | --- | --- |
| Field | Key words | Boolean Operator |
| Title | (Cancer* OR carcino* OR tumor* OR onco*) | AND |
| Title/Abstract | (colon* OR colorect*) | AND |
| Title/Abstract | (microbi* OR dysbi*) | AND |
| Title/Abstract | (chemo* OR immuno* OR car-t* OR radium* OR surgery* OR therapy*) | AND |
| Title/Abstract | (nutrition* OR diet* OR food* OR probio* OR prebio* OR symbio*) | NOT |
| Type of publication | (Biography OR Comment OR Duplicate publication OR Editorial OR Guideline OR Interview OR News OR Personal Narrative OR Published Erratum OR Retracted Publication OR Retraction of Publication OR Meta-Analysis OR Review OR Systematic Review) |  |
| Using words/prefixes with an asterisk opens the search to more results based on plurals and varying suffixes. |  |  |

The results of the different levels of the search can be found in the attached supplementary files.

#### Inclusion criteria

Only original articles were selected. Biographies, commentaries, duplicate publications, editorials, guides, interviews, news items, personal narratives, errata, retractions, reviews, systematic reviews and meta-analyses were excluded. In the next level of screening, only articles published in English that had mechanistic results and outcomes, at least *in vivo, were* selected by analysis of the abstract.

Publications that met the criteria above were downloaded and reviewed.

## 3. Results

### 3.1. Literature search

Figure 1 shows the flow diagram of the search and selection process. This diagram has been made by adapting the work to the guidelines for systematic searches recommended by Preferred Reporting Items for Systematic Reviews and Meta-Analyses (PRISMA).^114^ The Pubmed search yielded 202 results. The articles were screened for eligibility criteria. As a result of the first screening, a total of 108 articles were selected. After applying the next filtering on the type of results, a total of 62 articles remained for analysis. Of these 62 studies, 15 included human clinical trials (**Fig. 1**).

**Figure 1.**
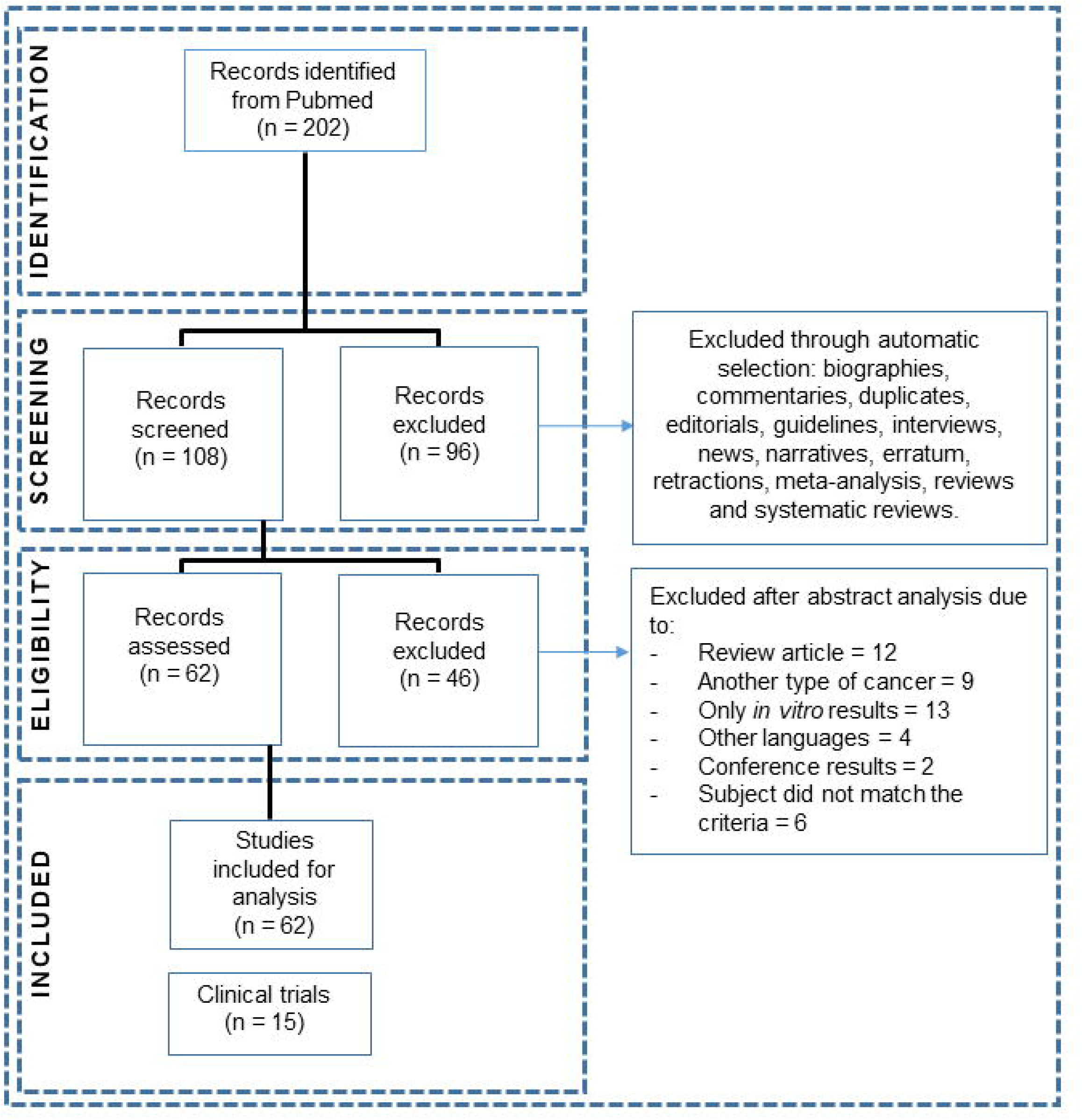
Flow chart of the search and selection process.

### 3.2. Characteristics of included results

Table 2 shows the characteristics of the included studies. The type of intervention performed, type of cancer, type of study, impact on the microbiota, the mechanism proposed by the authors, and its impact on cancer development are shown. Finally, the last column includes a more subjective criterion, namely the quality of the data analyzed on a scale ranging from very low = 1, low = 2, medium = 3, high = 4, very high = 5. This criterion has been assessed according to the experimental design, statistical analysis and strength of the mechanisms demonstrated. Figure 2 shows the distribution of the included studies.

**Table 2.**
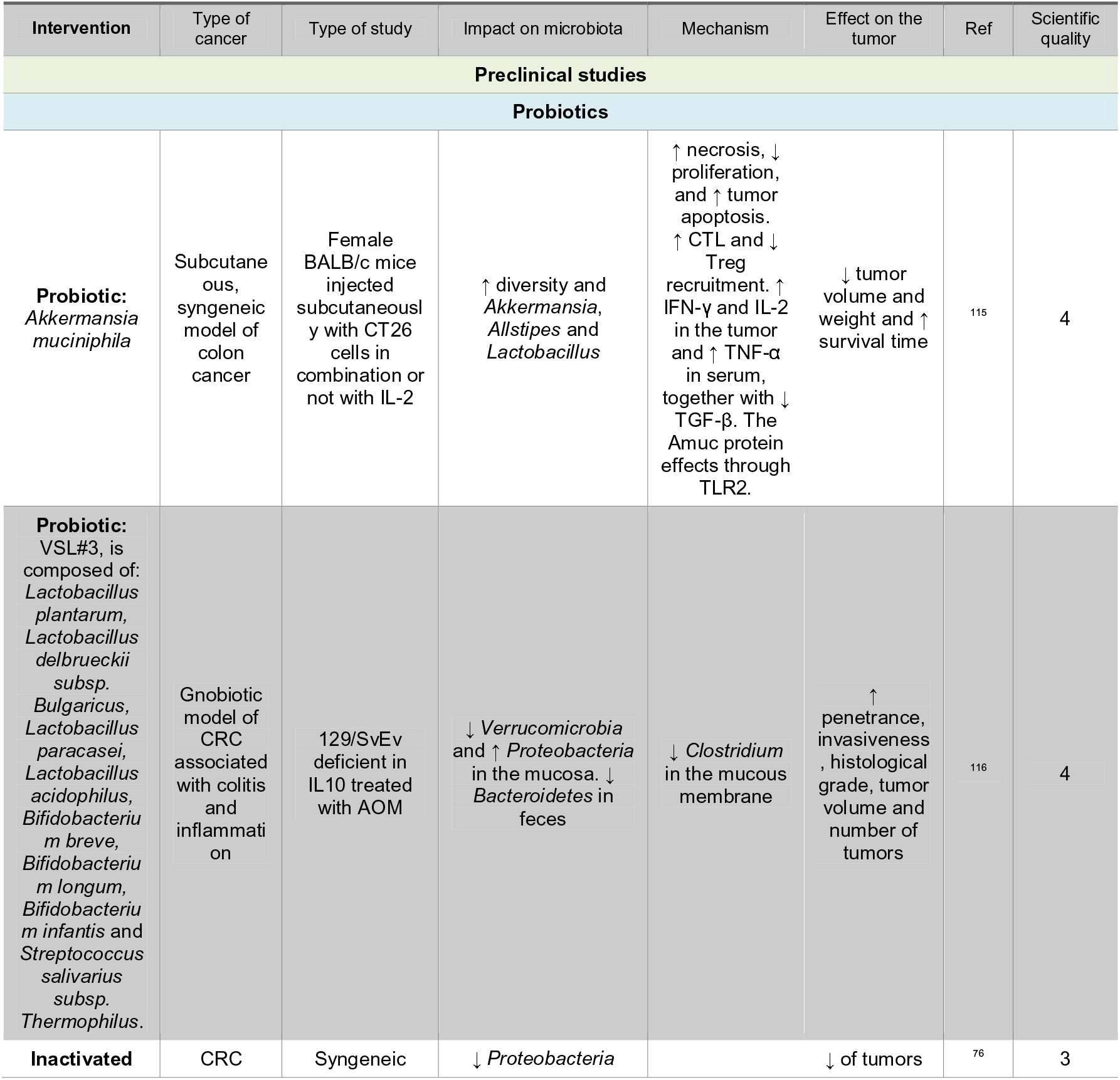
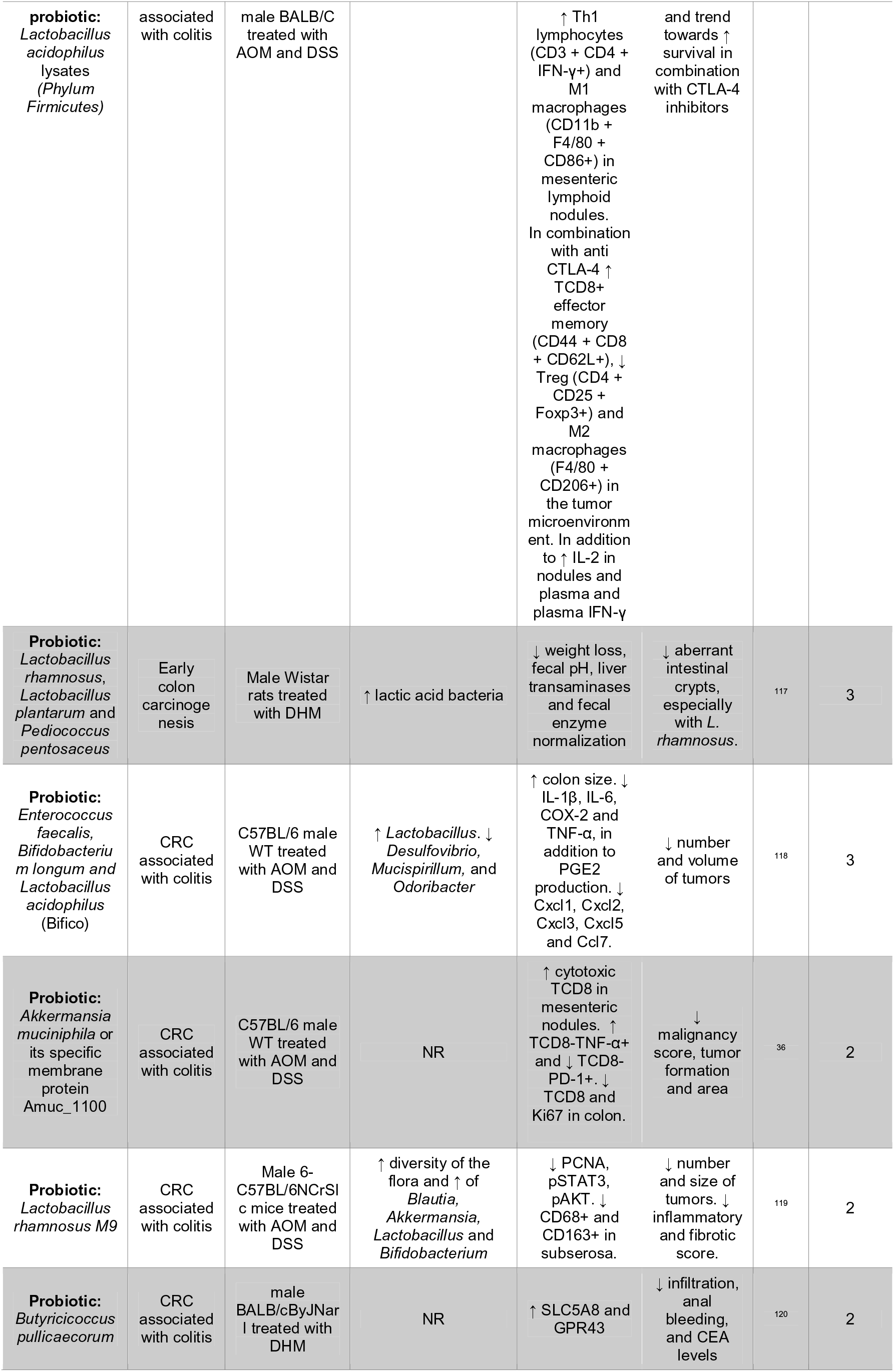
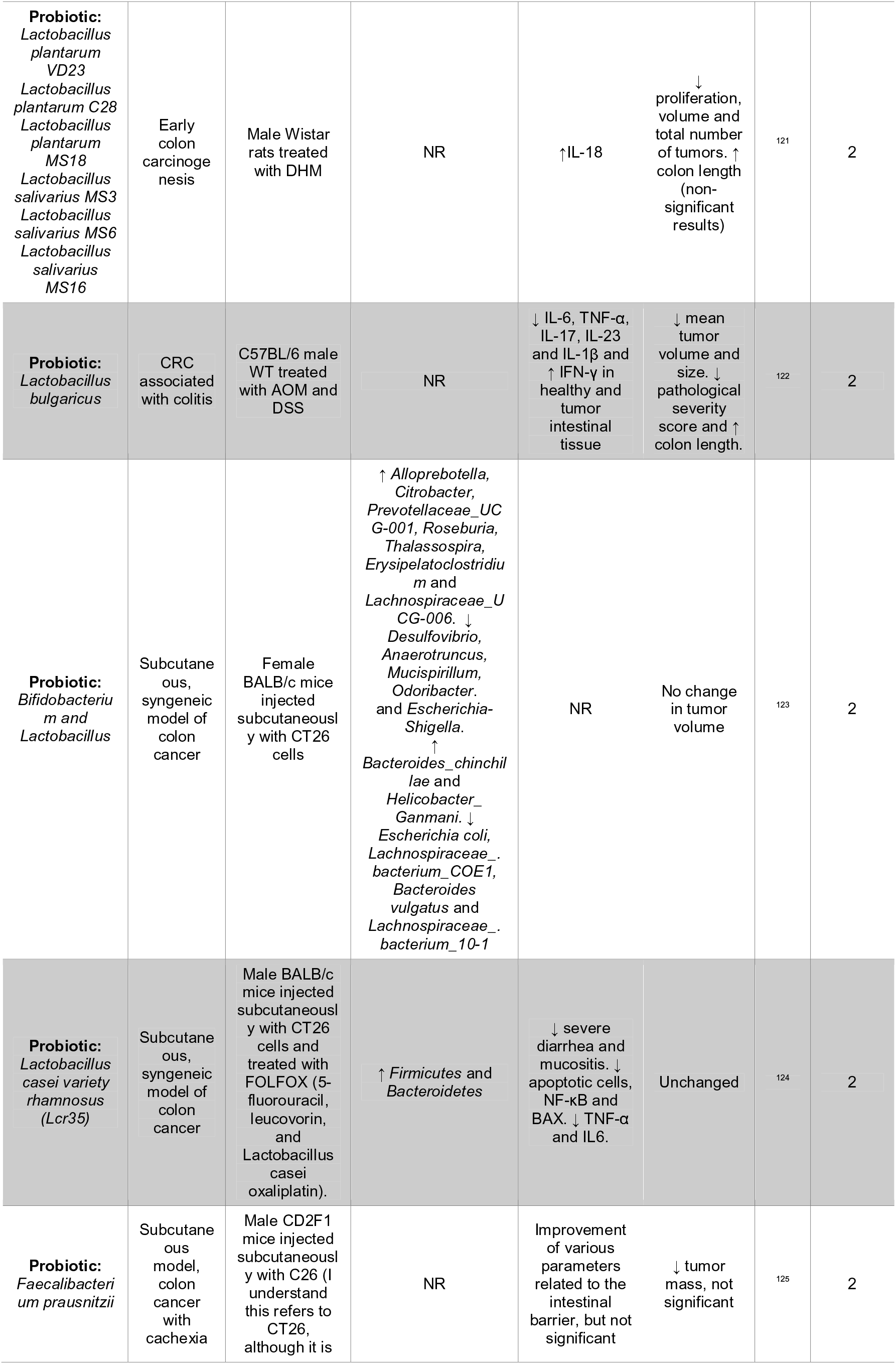
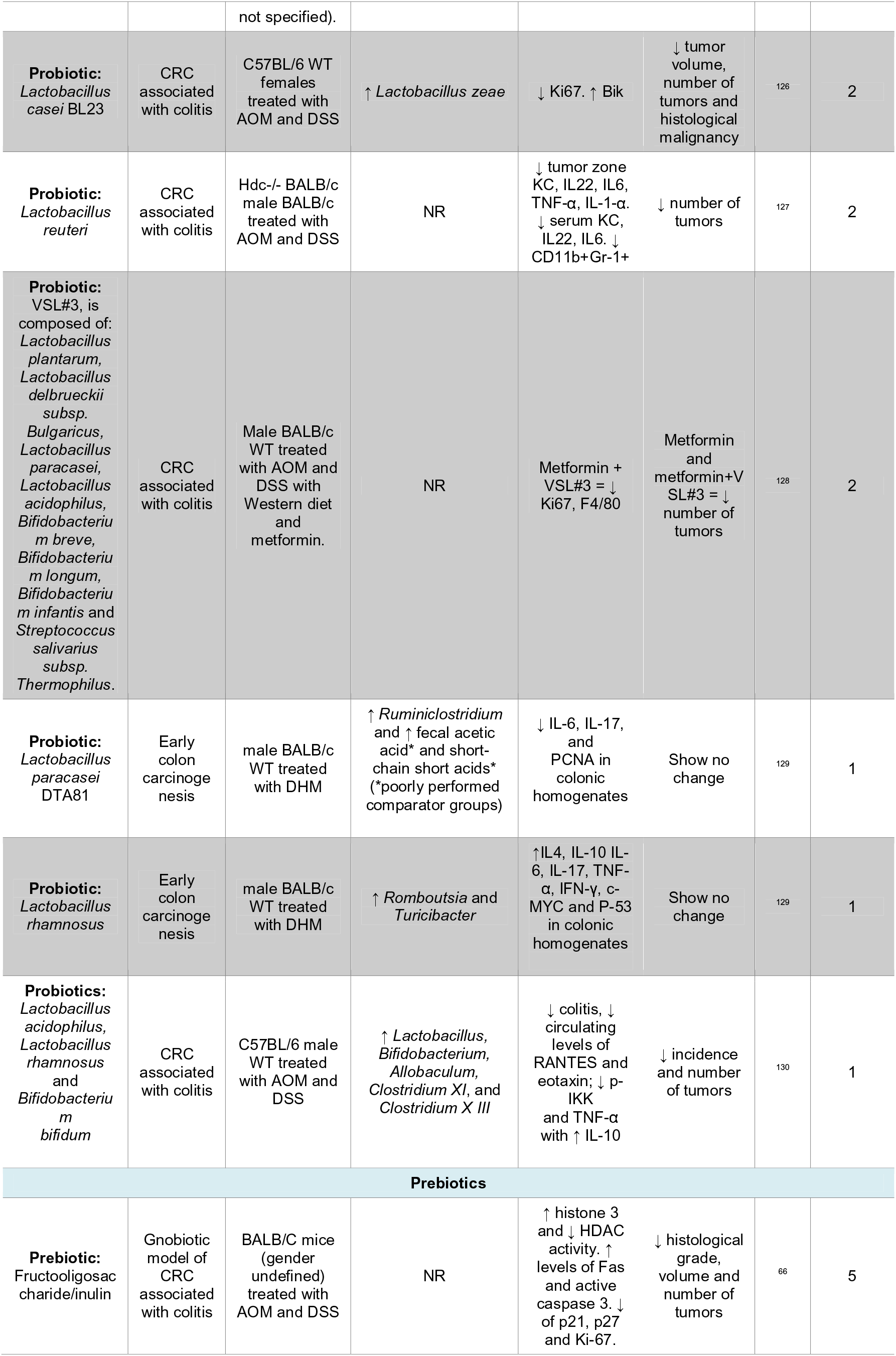
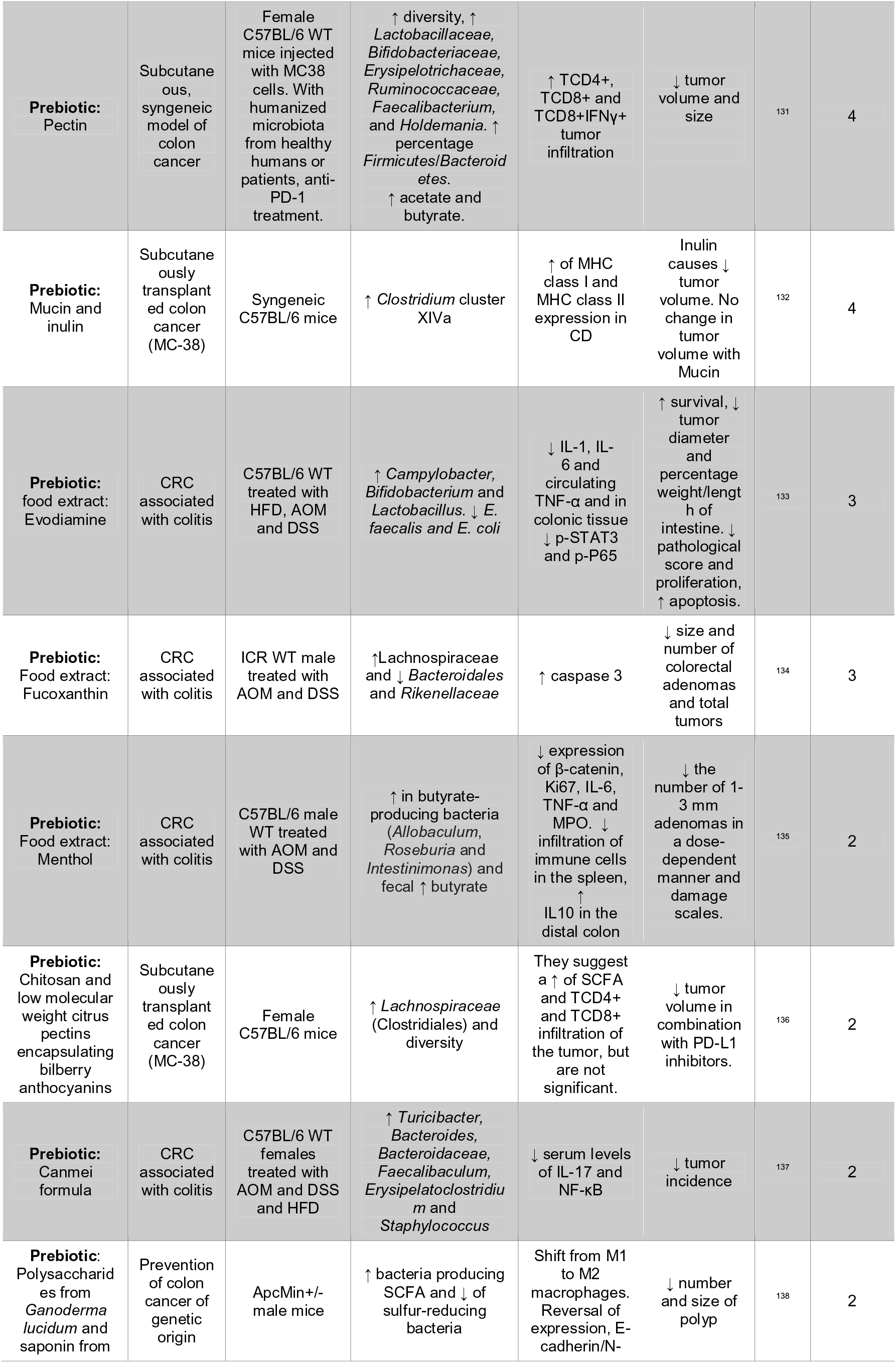
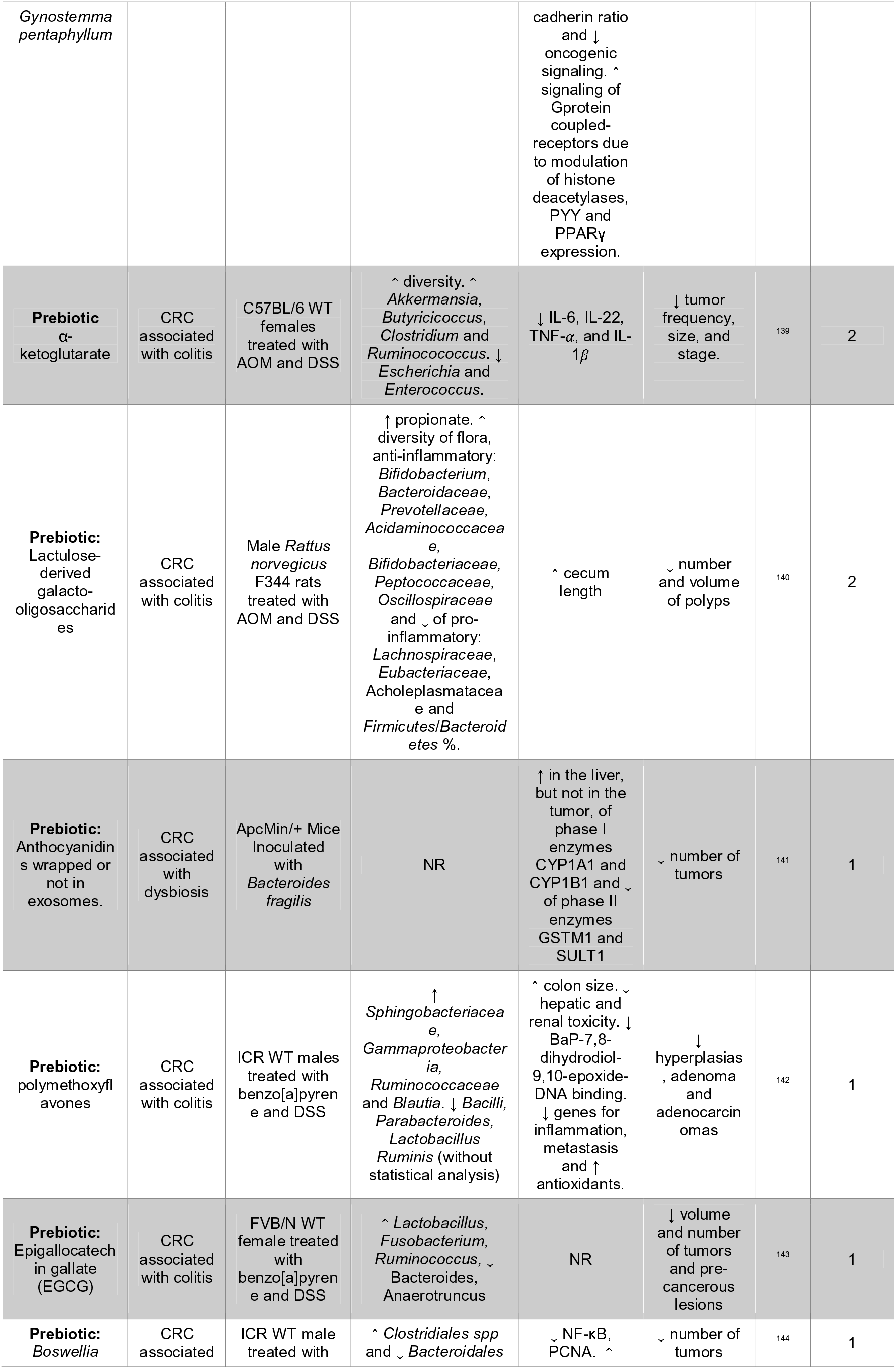
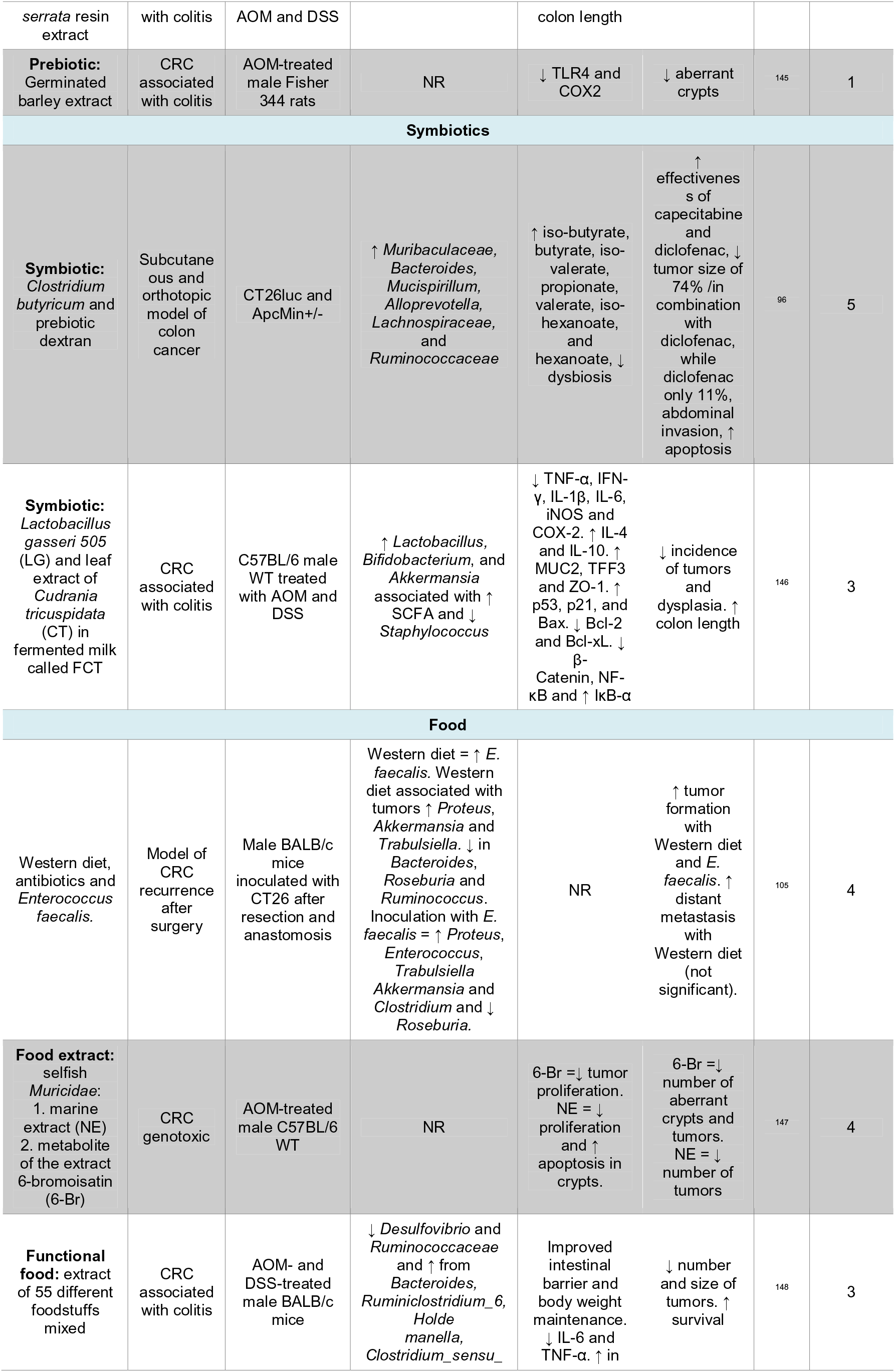
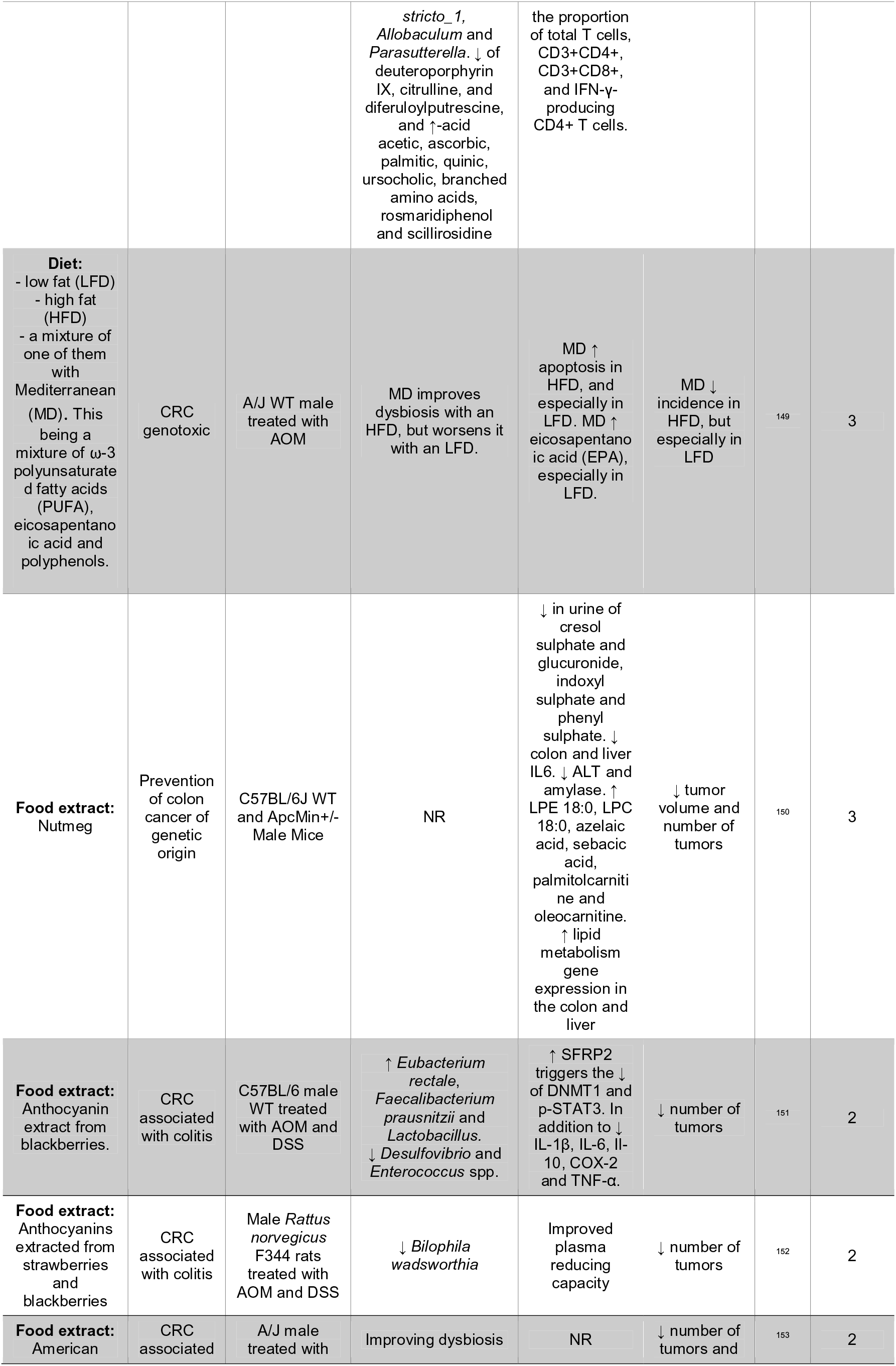
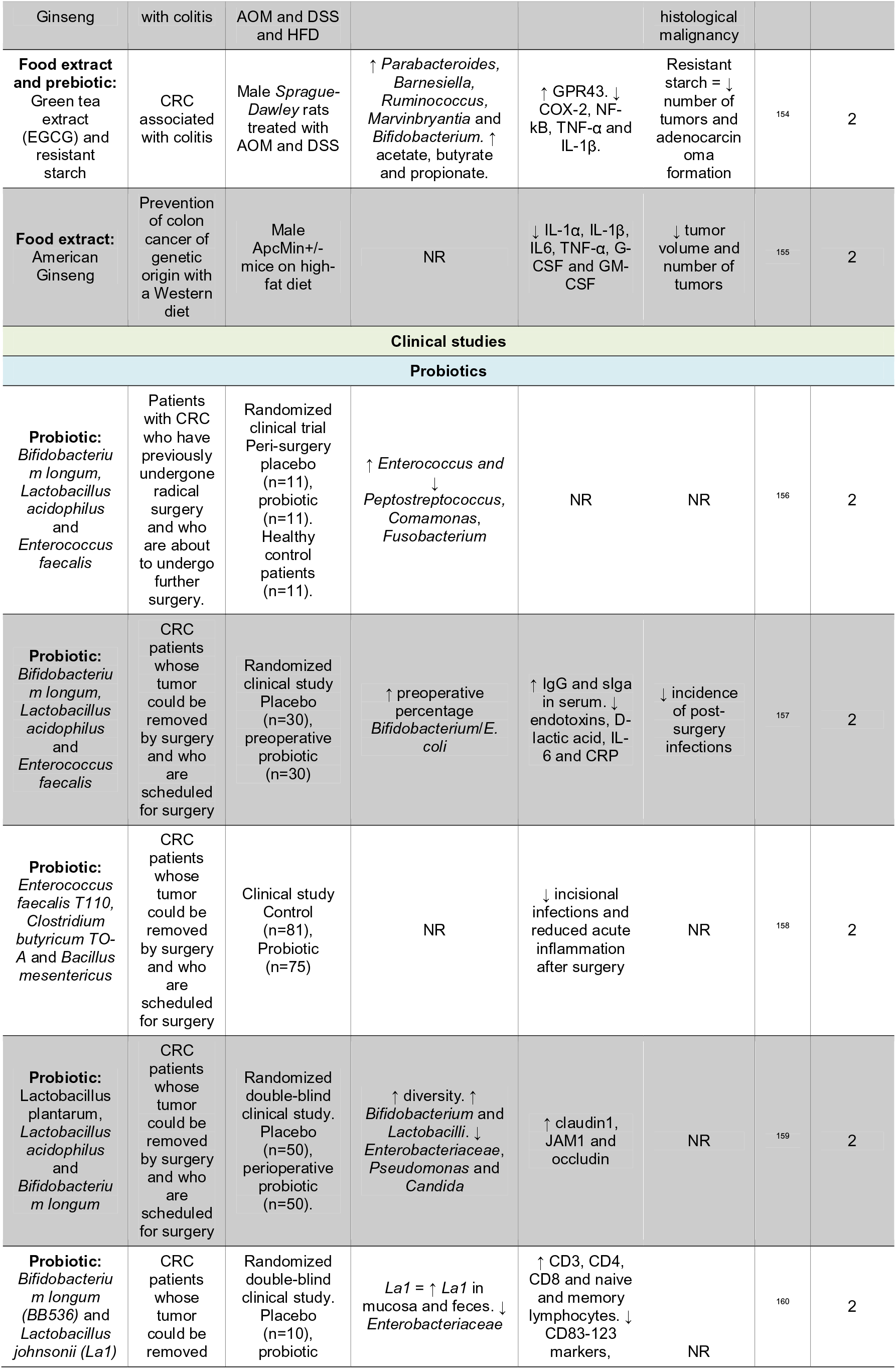
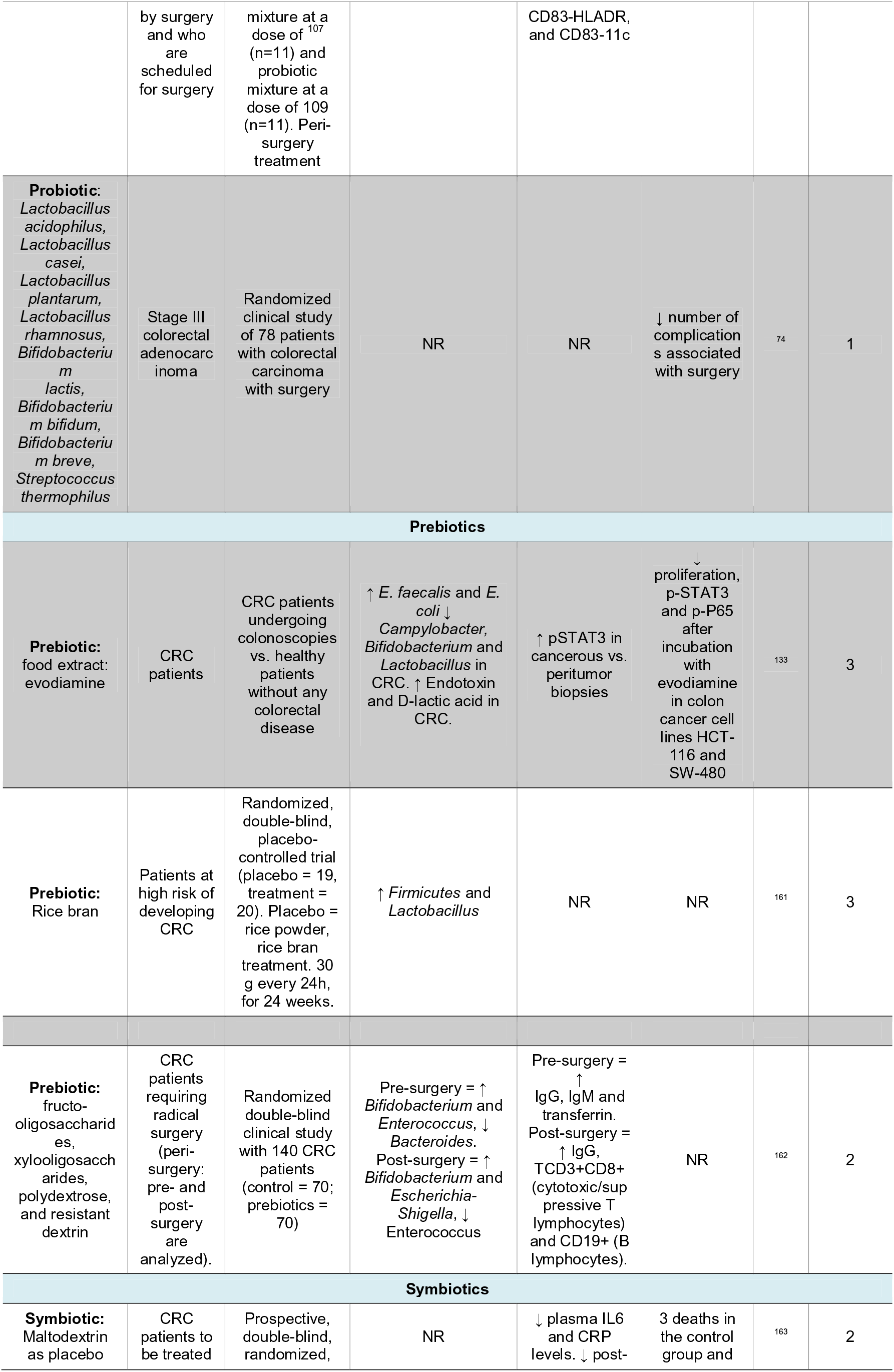
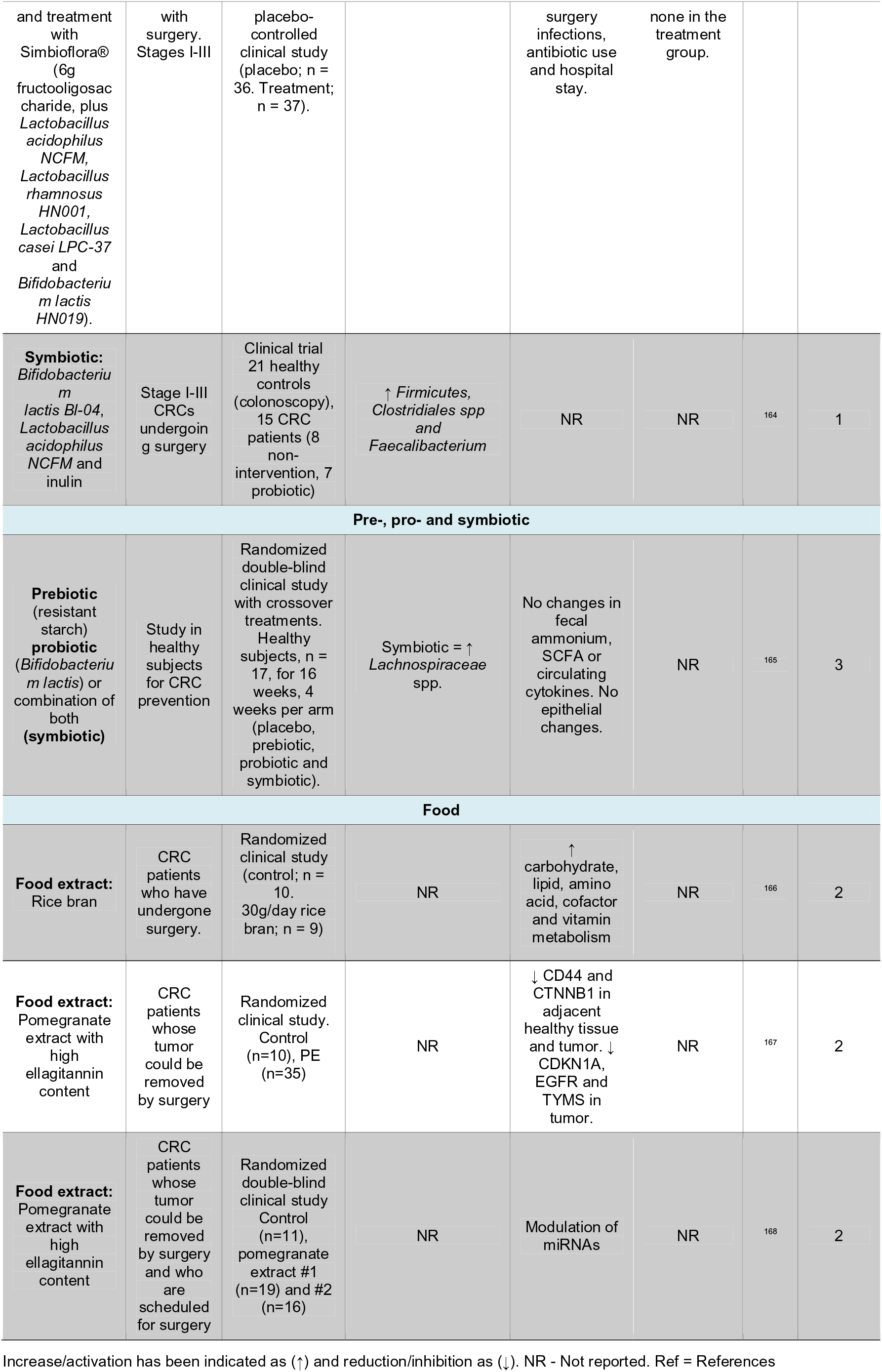
Summary of the results of the articles analyzed.

**Figure 2.**
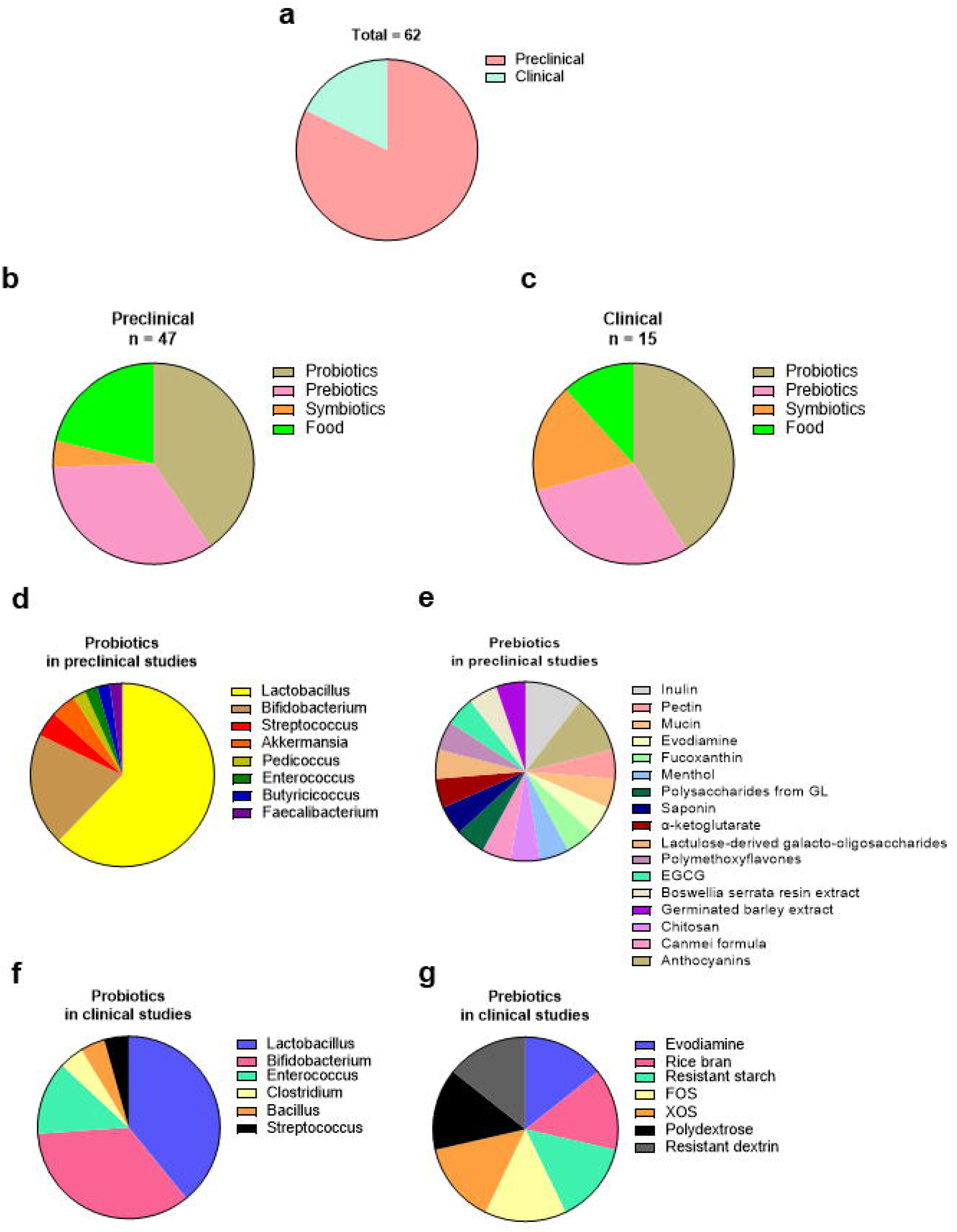
Distribution of the studies analyzed. **a.** Preclinical and clinical studies. **b and c.** Pre-, pro-, symbiotics and food in **b.** preclinical and **c.** clinical studies. **d.** Strains used as probiotics in preclinical studies. **e.** Prebiotics used in preclinical studies. **f.** Strains used as probiotics in clinical studies. **g.** Prebiotics used in clinical studies. * More than 1 pro- and prebiotic is used in some studies.

## 4. Discussion

A patient’s microbiota plays a key role in modulating the immune system as a natural defense against tumor development and during cancer treatment, especially in CRC.^20^ However, there is no defined therapeutic strategy for the microbiota in a CRC patient. To make the transition from clinical trials to patient treatment, it is necessary to review all published data on microbiota-related therapeutic approaches in CRC in a systematic way. The therapeutic modulators of the microbiome include primarily probiotics, prebiotics, symbiotics and nutrition.

### Probiotics

One of the most frequent probiotic combinations for the study of CRC is that of *Lactobacillus* and *Bifidobacterium*. In several studies, pre-inoculation with *Lactobacillus acidophilus* or a cocktail approved for commercial use in China, called Bifico, which consists of *Bifidobacterium longum*, *Lactobacillus acidophilus*, and *Enterococcus faecalis,* reduced colon tumor growth in preclinical models.^118, 169^ Bifico treatment also caused a decrease in *Desulfovibrio*, *Mucispirillum* and *Odoribacter*, associated with an enrichment in *Lactobacillus*.^118^ However, another study reported that a combination of *Bifidobacterium* and *Lactobacillus* was not able to improve 5-FU treatment, despite improving the flora composition.^123^

The mixture of three different strains of *Lactobacillus* and one of *Bifidobacterium* decreases the incidence and number of tumors in a preclinical model of CRC associated with colitis.^130^ VSL#3 is a commercial probiotic containing a mixture of different strains of *Lactobacillus, Bifidobacterium* and *Streptococcus.* In a murine model with a Western diet, treatment with VSL#3 and metformin decreases colonic tumor formation, through reduced cell proliferation and macrophage infiltration,.^128^ However, VSL#3 alone shows no protection against colonic tumor development in other studies.^116, 128^ In fact, in a model of colitis associated with CRC and inflammation, VSL#3 increases macroscopic lesions, as well as their invasiveness and histological grade.^116^ Therefore, it is important not to prescribe probiotics without scientific basis.

In addition, the *Lactobacillus* genus alone is probably the most widely tested probiotic in CRC (**Fig. 2 d&e**). A study conducted with *Lactobacillus casei* in a model of CRC associated with colitis, combined with FOLFOX, showed improvement in various inflammatory and microbiota parameters; however, it did not show reduction in tumor development.^124^ In this study, FOLFOX increases apoptosis in intestinal epithelial cells, while the probiotic inhibits cell death in healthy epithelial cells.^124^ Nonetheless, it is difficult to assess the effect of a given treatment in an animal model, but it is even more complicated in models that use injections of tumor cells in different (heterotopic) locations, as is the case in this study.^124^ Another study with the *L. casei* BL23 strain showed a total inhibition of the incidence of colorectal tumors in mice. This mechanism could be associated with increased expression of Bik, a proapoptotic gene, and a decrease in Ki67.^126^ *Lactobacillus reuteri* strain can also reduce colon tumors through the production of histamine, which is associated with a decrease in inflammatory markers at both local and systemic levels.^127^

Another well-studied probiotic is *Akkermansia muciniphila* (**Fig. 2 d**). Several studies support its potential use in CRC patients with promising results. Preclinical studies have shown that it is more efficient against CRC than IL-2 therapy. The combination of A. muciniphila and IL-2 was significantly more effective than their separate actions in reducing tumor volume and increasing survival.^115^ The effect of *A. muciniphila* appears to be related to a membrane protein called Amuc, which is recognized by Toll-like receptor 2 (TLR2). Cellular mechanisms and immune effects mediated by TLR2 lead to increased efficacy of IL-2 immune therapy.^115^

Therefore, it seems that probiotics, especially lactic acid bacteria, SCFA producers and *A. muciniphila* could be beneficial for patients diagnosed with CRC.

### Prebiotics

Many of the prebiotics used in the CRC literature are oligo- and polysaccharides (**Fig. 2 e & f**). Among them, pectin has shown beneficial effects such as SCFA production,^170^ reduction of ammonia,^171^ and improve glucose tolerance.^172^ Pectin is the fundamental metabolite of plant walls, and its fermentation serves as a source of energy for numerous microorganisms, producing metabolites with beneficial functions for our organism.^131, 173^ Furthermore, pectin is currently used in numerous food additives, so its study is of great interest.^131, 174^ In fact, it has been observed that pectin can be beneficial against several types of tumors such as CRC, breast and prostate cancer by halting tumor development.^174^ Its effect seems to be mediated through the increase of different SCFA-producing bacteria and in particular acetate, and the enhancement of the immune response through IFN-γ-producing TCD8+. In addition, a transplantation of fecal microbiota from PD-1 refractory patients showed enhanced anti-tumor response in PD-1 and pectin-treated mice.^131^

Galacto-oligosaccharides derived from lactulose, and mainly lactose, have been tested in preclinical models with promising results in terms of reducing tumor development and modulating the microbiota.^140^ Other well-studied oligosaccharides include fructo-oligosaccharides/inulin and mucin.^66, 132, 164^ Inulin has been shown to significantly reduce the volume of subcutaneously transplanted colon tumors, while mucin did not cause any significant change.^132^ In another study, inulin was reported to strongly inhibit tumor development through modification of the microbiota and butyrate production.^66^ This paper dissects the pathways leading to this effect, finding that butyrate is absorbed, but not oxidized due to the Warburg effect of cancer cells.^175^ However, healthy colonocytes do use butyrate as a major source of energy.^176^ In tumor cells, butyrate accumulates in the cell interior where it inhibits histone deacetylase activity, thus decreasing proliferation and increasing apoptosis.^66^ Studies with compounds derived from citrus peel, such as polymethoxyflavones, have shown positive effects in preventing the development of CRC. However, mechanisms such as autophagy, which the authors highlight as possible effector pathways for these compounds, are poorly analyzed and higher quality studies are needed to draw valid conclusions.^142^ Other prebiotics, such as green tea extract (EGCG), have been shown to reduce colonic tumors, but these studies are of low scientific quality (grading = 1 and 2) for the reasons mentioned above. In a rat model of colitis-associated cancer using EGCG and resistant starch, EGCG was shown to have no effect. Only resistant starch was able to decrease colonic tumor formation through a change in the microbial flora, increasing SCFA production, and decreasing inflammation.^154^

There is great interest among scientists in Asia, but particularly in China, in demonstrating the effects of different therapeutic approaches to traditional medicine through the microbiota.^135, 148^ For example, the Canmei formula contains herbs called *Mume sieb* and *Marci Hieronymi*, whose more than 41 active ingredients include DL-arginine, L-tyrosine, L(-)-carnitine, adenine, guanine or ambrosic acid. This formulation was shown to decrease the incidence of colonic tumors by reducing inflammation and improving dysbiosis.^137^ Another study using a *Boswellia serrata* resin extract has also demonstrated, in preclinical models, its ability to reduce colonic tumor formation.^144^ Many of these studies present positive results for the inhibition of CRC development. However, in general, they are of low quality (grading = 1 and 2) due to a lack of mechanistic and statistically robust data and therefore do not allow for reliable conclusions to be drawn.

### Symbiotics

One of the most interesting proposals in the literature is the possibility of using symbiotics as therapeutic options to increase the efficacy of available treatments. It has been observed that the use of *C. butyricum*, surrounded by dextran and bound to capecitabine or diclofenac, can increase the efficacy of these chemotherapeutics by >500%. This symbiotic decreases tumor invasiveness and also promotes a healthier and more diverse microbiota coupled with increased production of SCFA.^96^ This work is interesting for clinical studies as both dextran and *C. butyricum* are approved for use in food applications and showed no notable side effects.^96^ Another symbiotic studied is the combination of *Lactobacillus gasseri* 505 and *Cudrania tricuspidata* leaf extract in fermented milk. This symbiotic was shown to reduce tumor incidence and malignancy in a model of CRC-associated colitis. ^146^

Therefore, the use of symbiotics for improving gut microbiota in patients with CRC seems promising, although more studies are needed to clearly define which symbiotic should be used, the dose, the timing, and in which specific CRC patients.

### Healthy nutrition as a source of pre- and probiotics

Most, if not all pro-, pre- and symbiotics can be obtained naturally through a healthy diet. Prebiotics have been isolated from mushrooms,^138^ fruits,^142, 151^ dairy,^140^ rice,^161^ and green tea^138^ and probiotics from fruit skins,^117^ dairy,^122^ and fermented products.^146^

From infancy, a balanced, non-processed plant-based nutrition is the best way to have a proper gut microbiota. In fact, it has been shown that a diet high in soluble fiber is not able to reverse the erosion of intestinal mucus caused by a diet without fiber. However, the same diet, but high in intact plant fiber decreased mucus degradation, strengthening the intestinal barrier.^67^

Anthocyanins, compounds with a positive impact on the health of patients with CRC, are present in many foods such as tea, coffee, cocoa and red fruits. ^177^ In this regard, it has been shown that anthocyanins in blackberries can reduce the incidence of CRC, through diminished local inflammation and improved microbiota.^151^

Another food with interesting results is the marine extract of certain shellfish that may have anticarcinogenic effects. Using metabolomic analysis techniques, one can discern which compounds are toxic to tumor cells, opening the door to precision nutrition.^147, 178^ Algae-derived products such as fucoxanthin, a carotenoid present in high levels in wakame *(Undaria pinnatifida),* have also shown to decrease the number of adenocarcinomas and total tumors. This effect was also associated with an increase in the number of *Lachnospiraceae*.^134^ Spices have similarly attracted a great deal of interest for their health-promoting properties. For example, nutmeg can decrease the development of APC genetically engineered tumors by modulating the metabolism of the microbial flora and improving lipid metabolism.^150^ Numerous food extracts are also used in traditional Chinese medicine, including American ginseng. The use of this compound in a murine model prevented tumors and ameliorated dysbiosis induced by different murine models of CRC and high-fat diet.^153, 155^

Moving from specific foods to dietary patterns, the Mediterranean diet has been shown to have beneficial effects on multiple levels, specifically in the prevention of certain types of cancer.^179^ In the case of CRC, epidemiological studies show that adherence to a Mediterranean diet reduces the incidence of CRC by up to 11%.^180^ Likewise, the effect of mixing a Mediterranean diet with other types of diet (low-fat or high-fat Western-type diet) has been studied in models of cancer associated with colitis. This study showed that consumption of the Mediterranean diet, regardless of the diet with which it was mixed, decreased tumor incidence, and improved the microbiota, especially when accompanying a high-fat diet. ^149^

### Randomized clinical trials

At the highest level of scientific evidence are randomized clinical trials. Within this group, those conducted with lactic acid strains stand out. Among them, a study conducted with 78 patients analyzed the role of probiotics in reducing side effects after CRC surgery. Inclusion criteria were a diagnosis of stage III adenomatous CRC and that the patients were to undergo surgery. The probiotic group received post-operative treatment with a mixture of different strains of *Lactobacillus*, *Bifidobacterium,* and *Streptococcus thermophilus* for almost one year. The most important conclusion of this study was that probiotic treatment decreased the most frequent side effects of surgery, specifically paralytic ileus. However, limitations of this study include the low number of subjects included, the apparent lack of double-blinding, active placebo, and a weak statistical analysis.^74^ In another clinical trial, a probiotic consisting of *Bifidobacterium longum*, *Lactobacillus acidophilus* and *Enterococcus faecalis* was tested in CRC patients undergoing surgery. It was shown that, in addition to improving the diversity of the microbial flora, the probiotic mixture was able to decrease the levels of *Fusobacterium*, a bacterium associated with the development of CRC, by >500%.^156^ In addition, its use decreased post-operative infections.^157^ Another similar probiotic mixture consisting of *Lactobacillus plantarum*, *Lactobacillus acidophilus* and *Bifidobacterium longum* reduced the risk of postoperative infection and improved the intestinal epithelial barrier, resulting in a lower incidence of diarrhea, abdominal pain, and fever.^159^ A study using a mixture of *Bifidobacterium longum* (BB536) and *Lactobacillus johnsonii* (La1) showed that only the latter (La1) colonized the gut, reducing the number of pathogenic microorganisms and reducing colonic inflammation in CRC patients undergoing surgery. This result shows that adding more probiotics will not have a greater effect and it is important to study each strain individually and its combination before it can be recommended to patients.^160^

In another clinical trial, a probiotic mixture consisting of *Enterococcus, Clostridium* and *Bacillus* was used in patients undergoing CRC surgery to discern whether their consumption could ameliorate the adverse effects of surgery. It was found that peri-surgical use of this mixture significantly reduced incisional infections and decreased the acute inflammatory response to surgery.^158^ However, in this study, no randomization was performed, and patients were included in one group (control) or the other (probiotic) according to the date of surgery, with the control group (2009-2011) prior to the probiotic group (2011-2013). This does not allow for a clear conclusion on the effect of the probiotic, because multiple factors may have changed during those two years, including greater experience of the surgical team or better technical means. In this regard, the authors describe that a higher number of laparotomies were performed in the probiotic group, and that performing or not performing laparotomy is a key factor related to the outcome of the surgery.^158^

As far as prebiotics are concerned, rice bran is of great interest due to its large commercial use. Numerous studies show the benefit of whole grain consumption in the prevention of CRC.^181^ In the case of rice bran, various phytochemicals in its composition have been associated with a decrease in the risk of cancer development, in particular CRC.^182^ Its consumption increases levels of *Firmicutes* and *Lactobacillus*^161^ and produces numerous metabolic modifications that could be related to a decrease in the risk of developing CRC.^166^

In another clinical trial with prebiotics in CRC patients, a mixture of different non-digestible oligosaccharides was used for 7 days before patients underwent radical surgery to remove the tumor. Both groups received the same nutrition (provided by the hospital) in the 7 days before and after surgery (parenteral nutrition). Surgery decreased the diversity of the microbiota in both groups, underlining the impact of these interventions on gut health. Interestingly, a principal component analysis of microbiome diversity was able to unequivocally identify samples from postoperative patients who had received prebiotics. Furthermore, the use of these prebiotics improved systemic immune function, confirming the positive effect that microbiota can have on immune response. An important limitation of this study is that the randomization did not specify the stage or type of tumor.^162^

Symbiotics have also been tested in randomized clinical trials. A commercially named symbiotic, simbioflora®, containing fructooligosaccharides, 3 strains of *Lactobacillus,* and *Bifidobacterium lactis* showed promising results in CRC patients undergoing surgery. The use of this symbiotic significantly reduced inflammation and postoperative infections.^163^ However, this is a small study of patients with cancer at different stages, so the data should be analyzed with caution. In another study, a mixture of *Bifidobacterium*, *Lactobacillus* and inulin strains was used in stage I-III patients undergoing surgery. This treatment resulted in an improvement in the composition of the microbial community, especially the SCFA-producing bacterium. However, this study does not define blind randomization of patients and there is no placebo, so the data should be also analyzed with caution.^164^ Finally, a randomized clinical study was conducted in 20 healthy subjects with backcrossed arms with the aim of preventing CRC. This study of 4 weeks duration per intervention (switching every 4 weeks to another arm until each group completed all 4) was done with 4 different arms: placebo, *Bifidobacterium lactis,* resistant starch and their combination (*Bifidobacterium lactis* and resistant starch). It was shown that in healthy subjects the combination of *Bifidobacterium lactis* and resistant starch increased the levels of *Lachnospiraceae spp.* However, the other parameters analyzed, such as stool mass, pH or ammonium, SCFA, circulating cytokines, and markers of colonic epithelial health, did not change.^165^

The use of pomegranate extract in patients undergoing surgical tumor resection has also been investigated. Pomegranate extract was shown to produce significant changes in the expression of numerous genes and microRNAs related to tumor development. These results underline the crucial role of diet in modifying gene expression and, therefore, in the etiology of disease.^167, 168^

## 5. Conclusion

We can summarize the conclusions of this work regarding the modification of the microbiota in the CRC patient as follows:

1. **Antibiotic use** is one of the **major deleterious factors** for the prognosis of treatment efficacy, **due to its** harmful effect on the microbiota.
2. **Prebiotics** such as inulin and resistant starch can support the development of a healthy microbiota.
3. **Probiotics**, mainly based on **SCFA-producing lactic acid strains,** show a **favorable profile** in preclinical and clinical studies. However, not all strains are equally favorable and are therefore <u>not recommended for use in patients without individualized study of the patient and the probiotic.</u>
4. A **healthy** and varied **nutrition**, consisting mainly of unprocessed **plant-based foods**, seems to be one of the **best sources** to maintain a **homeostatic microbiota**.

Finally, after a detailed analysis of the literature, in a **healthy person**, <u>there is no need</u> to recommend the use of <u>pre-, pro- or symbiotics</u> in addition to a **healthy diet**. However, CRC patients often have <u>severe dysbiosis</u> due to their own disease, and broad-spectrum antibiotic use. In these patients, apart from a **healthy diet**, or if a healthy diet cannot be established, <u>carefully chosen pre-, pro- and/or symbiotics may be beneficial to</u> restore a beneficial microbiota and support the efficacy of hospital treatments more effectively.

## Data Availability

There are not restrictions on data availability in this manuscript.

## Acknowledgements

We thank Dr. Alberto Canfran-Duque and Noemi Rotllan for their for critical reading of the manuscript. We apologize to those whose work could not be cited owing to space limitations.

## Authors Contributions

JMM designed and performed the systematic search, wrote and edited the manuscript. SBE contributed to the editing and interpretation of the manuscript. All authors read and edited the final draft of the manuscript.

## Competing Financial Interest

The authors declare no conflicts of interest

Correspondence and requests for materials should be addressed to.

## Data availability

There are not restrictions on data availability in this manuscript.

## Abbreviations

CRC: colorectal cancer
ROS: reactive oxygen species
RNS: reactive nitrogen species
MSI: Microsatellite instability
SCFA: Short chain fatty acids
LPS: Lipopolysaccharide
dMMR: deficient DNA mismatch repair
AOM: Azoxymethane
DSS: Dextran sodium sulfate
TNF-α: tumor necrosis factor alpha
MPO: Myeloperoxidase
PCNA: Proliferation cellular nuclear antigen
SLC5A8: Solute Carrier Family 5 Member 8
GPR43: G protein-coupled receptor 43
CEA: Carcinoembryonic antigen
APC: Adenomatous polyposis coli
DHM: 1,2-dimethyl hydrazine
TGF-β: Transforming growth factor beta
TLR2: toll-like receptor 2
DC: dendritic cells
PD-1: Programmed cell death protein 1
PDL-1: Programmed cell death protein ligand 1
CTLA-4: Cytotoxic T-Lymphocyte Associated Protein 4
CYP: Cytochrome P450 Family
GSTM1: Glutathione S-Transferase Mu 1
SULT1: Sulfotransferase Family-1
PYY: Peptide tyrosine
PPARγ: peroxisome proliferator activated receptor gamma
6,BR: 6-bromoisatin
NE: marine extract
MD: Mediterranean diet
LFD: low fat diet
HFD: high fat diet
EPA: Eicosapentanoic acid
PUFA: polyunsaturated fatty acids
CRP: C reactive protein
SFRP2: Secreted Frizzled Related Protein 2
DNMT1: DNA Methyltransferase 1
COX-2: Cyclooxygenase-2
PGE_2_: Prostaglandin E2
Cxcl: C-X-C Motif Chemokine Ligand
Bik: BCL2 Interacting Killer
KC: Keratinocytes-derived chemokine
Gr-1: Glucocorticoid Receptor-1
CTNNB1: Beta catenin 1
EGFR: Epidermal growth factor receptor
TYMS: Thymidylate Synthetase
LPE: Lisophosphatidiletanolamine
LPC: Lisophosphatidylcholine
G-CSF: Granulocyte Colony-Stimulating Factor
GM-CSF: Granulocyte Macrophage Colony-Stimulating Factor
HDAC: Histone Deacetylase
JAM1: F11 Receptor
5-FU: 5-florouracil
EGCG: epigallocatechin-3-gallate

## Bibliography

1. Henrikson, N.B. et al. Family history and the natural history of colorectal cancer: systematic review. Genet Med 17, 702–712 (2015).

2. Czene, K., Lichtenstein, P. & Hemminki, K. Environmental and heritable causes of cancer among 9.6 million individuals in the Swedish Family-Cancer Database. Int J Cancer 99, 260–266 (2002).

3. Lichtenstein, P. et al. Environmental and heritable factors in the causation of cancer--analyses of cohorts of twins from Sweden, Denmark, and Finland. N Engl J Med 343, 78–85 (2000).

4. Jiao, S. et al. Estimating the heritability of colorectal cancer. Hum Mol Genet 23, 3898–3905 (2014).

5. Syngal, S. et al. ACG clinical guideline: Genetic testing and management of hereditary gastrointestinal cancer syndromes. Am J Gastroenterol 110, 223–262; quiz 263 (2015).

6. Dekker, E., Tanis, P.J., Vleugels, J.L.A., Kasi, P.M. & Wallace, M.B. Colorectal cancer. Lancet 394, 1467–1480 (2019).

7. Nishida, A. et al. Gut microbiota in the pathogenesis of inflammatory bowel disease. Clin J Gastroenterol 11, 1–10 (2018).

8. Murphy, N. et al. Lifestyle and dietary environmental factors in colorectal cancer susceptibility. Mol Aspects Med 69, 2–9 (2019).

9. Crosnier, C., Stamataki, D. & Lewis, J. Organizing cell renewal in the intestine: stem cells, signals and combinatorial control. Nat Rev Genet 7, 349–359 (2006).

10. (IARC Working Group on the Evaluation of Carcinogenic Risks to Humans. Lyon (FR): International Agency for Research on Cancer; 2010. (IARC Monographs on the Evaluation of Carcinogenic Risks to Humans, No. 92.); 2010).

11. Mottram, D.S., Wedzicha, B.L. & Dodson, A.T. Acrylamide is formed in the Maillard reaction. Nature 419, 448–449 (2002).

12. (IARC Working Group on the Evaluation of Carcinogenic Risks to Humans. Chemical Agents and Related Occupations. Lyon (FR): International Agency for Research on Cancer; 2012. (IARC Monographs on the Evaluation of Carcinogenic Risks to Humans, No. 100F.) 2012).

13. Diggs, D.L. et al. Polycyclic aromatic hydrocarbons and digestive tract cancers: a perspective. J Environ Sci Health C Environ Carcinog Ecotoxicol Rev 29, 324–357 (2011).

14. Tabatabaei, S.M., Heyworth, J.S., Knuiman, M.W. & Fritschi, L. Dietary benzo[a]pyrene intake from meat and the risk of colorectal cancer. Cancer Epidemiol Biomarkers Prev 19, 3182–3184 (2010).

15. Coussens, L.M. & Werb, Z. Inflammation and cancer. Nature 420, 860–867 (2002).

16. Medema, J.P. Cancer stem cells: the challenges ahead. Nat Cell Biol 15, 338–344 (2013).

17. Cancer Genome Atlas, N. Comprehensive molecular characterization of human colon and rectal cancer. Nature 487, 330–337 (2012).

18. Stappenbeck, T.S. & Virgin, H.W. Accounting for reciprocal host-microbiome interactions in experimental science. Nature 534, 191–199 (2016).

19. Zitvogel, L., Ma, Y., Raoult, D., Kroemer, G. & Gajewski, T.F. The microbiome in cancer immunotherapy: Diagnostic tools and therapeutic strategies. Science 359, 1366–1370 (2018).

20. Thursby, E. & Juge, N. Introduction to the human gut microbiota. Biochem J 474, 1823–1836 (2017).

21. Claesson, M.J. et al. Gut microbiota composition correlates with diet and health in the elderly. Nature 488, 178–184 (2012).

22. Wang, T. et al. Structural segregation of gut microbiota between colorectal cancer patients and healthy volunteers. ISME J 6, 320–329 (2012).

23. Lin, Y. et al. NMR-based fecal metabolomics fingerprinting as predictors of earlier diagnosis in patients with colorectal cancer. Oncotarget 7, 29454–29464 (2016).

24. Chen, J. & Vitetta, L. Inflammation-Modulating Effect of Butyrate in the Prevention of Colon Cancer by Dietary Fiber. Clin Colorectal Cancer 17, e541–e544 (2018).

25. Zeng, H., Safratowich, B.D., Wang, T.T.Y., Hamlin, S.K. & Johnson, L.K. Butyrate Inhibits Deoxycholic-Acid-Resistant Colonic Cell Proliferation via Cell Cycle Arrest and Apoptosis: A Potential Pathway Linking Dietary Fiber to Cancer Prevention. Mol Nutr Food Res 64, e1901014 (2020).

26. Tan, J. et al. The role of short-chain fatty acids in health and disease. Adv Immunol 121, 91–119 (2014).

27. Ganapathy, V., Thangaraju, M., Prasad, P.D., Martin, P.M. & Singh, N. Transporters and receptors for short-chain fatty acids as the molecular link between colonic bacteria and the host. Curr Opin Pharmacol 13, 869–874 (2013).

28. Zhao, Y. et al. GPR43 mediates microbiota metabolite SCFA regulation of antimicrobial peptide expression in intestinal epithelial cells via activation of mTOR and STAT3. Mucosal Immunol 11, 752–762 (2018).

29. Madara, J.L. & Pappenheimer, J.R. Structural basis for physiological regulation of paracellular pathways in intestinal epithelia. J Membr Biol 100, 149–164 (1987).

30. Eeckhaut, V. et al. Butyricicoccus pullicaecorum in inflammatory bowel disease. Gut 62, 1745–1752 (2013).

31. Kang, S. et al. Dysbiosis of fecal microbiota in Crohn’s disease patients as revealed by a custom phylogenetic microarray. Inflamm Bowel Dis 16, 2034–2042 (2010).

32. Takaishi, H. et al. Imbalance in intestinal microflora constitution could be involved in the pathogenesis of inflammatory bowel disease. Int J Med Microbiol 298, 463–472 (2008).

33. Karlsson, C.L. et al. The microbiota of the gut in preschool children with normal and excessive body weight. Obesity (Silver Spring) 20, 2257–2261 (2012).

34. Crovesy, L., Masterson, D. & Rosado, E.L. Profile of the gut microbiota of adults with obesity: a systematic review. Eur J Clin Nutr 74, 1251–1262 (2020).

35. Vigsnaes, L.K., Brynskov, J., Steenholdt, C., Wilcks, A. & Licht, T.R. Gram-negative bacteria account for main differences between faecal microbiota from patients with ulcerative colitis and healthy controls. Benef Microbes 3, 287–297 (2012).

36. Wang, L. et al. A purified membrane protein from Akkermansia muciniphila or the pasteurised bacterium blunts colitis associated tumourigenesis by modulation of CD8(+) T cells in mice. Gut 69, 1988–1997 (2020).

37. Ottman, N. et al. Genome-Scale Model and Omics Analysis of Metabolic Capacities of Akkermansia muciniphila Reveal a Preferential Mucin-Degrading Lifestyle. Appl Environ Microbiol 83 (2017).

38. Wang, G. et al. Role of SCFAs in gut microbiome and glycolysis for colorectal cancer therapy. J Cell Physiol 234, 17023–17049 (2019).

39. Petersen, C. & Round, J.L. Defining dysbiosis and its influence on host immunity and disease. Cell Microbiol 16, 1024–1033 (2014).

40. Wong, S.H. & Yu, J. Gut microbiota in colorectal cancer: mechanisms of action and clinical applications. Nat Rev Gastroenterol Hepatol 16, 690–704 (2019).

41. Peters, B.A. et al. The gut microbiota in conventional and serrated precursors of colorectal cancer. Microbiome 4, 69 (2016).

42. Dethlefsen, L., Huse, S., Sogin, M.L. & Relman, D.A. The pervasive effects of an antibiotic on the human gut microbiota, as revealed by deep 16S rRNA sequencing. PLoS Biol 6, e280 (2008).

43. Henao-Mejia, J. et al. Inflammasome-mediated dysbiosis regulates progression of NAFLD and obesity. Nature 482, 179–185 (2012).

44. Newton, D.F., Cummings, J.H., Macfarlane, S. & Macfarlane, G.T. Growth of a human intestinal Desulfovibrio desulfuricans in continuous cultures containing defined populations of saccharolytic and amino acid fermenting bacteria. J Appl Microbiol 85, 372–380 (1998).

45. Carbonero, F., Benefiel, A.C. & Gaskins, H.R. Contributions of the microbial hydrogen economy to colonic homeostasis. Nat Rev Gastroenterol Hepatol 9, 504–518 (2012).

46. Roediger, W.E., Duncan, A., Kapaniris, O. & Millard, S. Reducing sulfur compounds of the colon impair colonocyte nutrition: implications for ulcerative colitis. Gastroenterology 104, 802–809 (1993).

47. Suarez, F., Furne, J., Springfield, J. & Levitt, M. Production and elimination of sulfur-containing gases in the rat colon. Am J Physiol 274, G727–733 (1998).

48. Cai, W.J., Wang, M.J., Ju, L.H., Wang, C. & Zhu, Y.C. Hydrogen sulfide induces human colon cancer cell proliferation: role of Akt, ERK and p21. Cell Biol Int 34, 565–572 (2010).

49. Deplancke, B. & Gaskins, H.R. Hydrogen sulfide induces serum-independent cell cycle entry in nontransformed rat intestinal epithelial cells. FASEB J 17, 1310–1312 (2003).

50. Attene-Ramos, M.S., Wagner, E.D., Plewa, M.J. & Gaskins, H.R. Evidence that hydrogen sulfide is a genotoxic agent. Mol Cancer Res 4, 9–14 (2006).

51. Kanazawa, K. et al. Factors influencing the development of sigmoid colon cancer. Bacteriologic and biochemical studies. Cancer 77, 1701–1706 (1996).

52. Kostic, A.D. et al. Genomic analysis identifies association of Fusobacterium with colorectal carcinoma. Genome Res 22, 292–298 (2012).

53. Castellarin, M. et al. Fusobacterium nucleatum infection is prevalent in human colorectal carcinoma. Genome Res 22, 299–306 (2012).

54. Yang, Y. et al. Fusobacterium nucleatum Increases Proliferation of Colorectal Cancer Cells and Tumor Development in Mice by Activating Toll-Like Receptor 4 Signaling to Nuclear Factor-kappaB, and Up-regulating Expression of MicroRNA-21. Gastroenterology 152, 851–866 e824 (2017).

55. Yu, T. et al. Fusobacterium nucleatum Promotes Chemoresistance to Colorectal Cancer by Modulating Autophagy. Cell 170, 548–563 e516 (2017).

56. Campisciano, G. et al. The Obesity-Related Gut Bacterial and Viral Dysbiosis Can Impact the Risk of Colon Cancer Development. Microorganisms 8 (2020).

57. Feng, Q. et al. Gut microbiome development along the colorectal adenoma-carcinoma sequence. Nat Commun 6, 6528 (2015).

58. Arthur, J.C. et al. Intestinal inflammation targets cancer-inducing activity of the microbiota. Science 338, 120–123 (2012).

59. Garrett, W.S. The gut microbiota and colon cancer. Science 364, 1133–1135 (2019).

60. Dejea, C.M. et al. Patients with familial adenomatous polyposis harbor colonic biofilms containing tumorigenic bacteria. Science 359, 592–597 (2018).

61. Zhang, G., Svenungsson, B., Karnell, A. & Weintraub, A. Prevalence of enterotoxigenic Bacteroides fragilis in adult patients with diarrhea and healthy controls. Clin Infect Dis 29, 590–594 (1999).

62. Turnbaugh, P.J. et al. A core gut microbiome in obese and lean twins. Nature 457, 480–484 (2009).

63. Turnbaugh, P.J. et al. The effect of diet on the human gut microbiome: a metagenomic analysis in humanized gnotobiotic mice. Sci Transl Med 1, 6ra14 (2009).

64. Carmody, R.N. et al. Diet dominates host genotype in shaping the murine gut microbiota. Cell Host Microbe 17, 72–84 (2015).

65. De Filippis, F. et al. High-level adherence to a Mediterranean diet beneficially impacts the gut microbiota and associated metabolome. Gut 65, 1812–1821 (2016).

66. Donohoe, D.R. et al. A gnotobiotic mouse model demonstrates that dietary fiber protects against colorectal tumorigenesis in a microbiota- and butyrate-dependent manner. Cancer Discov 4, 1387–1397 (2014).

67. Desai, M.S. et al. A Dietary Fiber-Deprived Gut Microbiota Degrades the Colonic Mucus Barrier and Enhances Pathogen Susceptibility. Cell 167, 1339–1353 e1321 (2016).

68. Halmos, E.P., Power, V.A., Shepherd, S.J., Gibson, P.R. & Muir, J.G. A diet low in FODMAPs reduces symptoms of irritable bowel syndrome. Gastroenterology 146, 67–75 e65 (2014).

69. Sanders, M.E., Merenstein, D.J., Reid, G., Gibson, G.R. & Rastall, R.A. Probiotics and prebiotics in intestinal health and disease: from biology to the clinic. Nat Rev Gastroenterol Hepatol 16, 605–616 (2019).

70. Nami, Y., Haghshenas, B., Haghshenas, M., Abdullah, N. & Yari Khosroushahi, A. The Prophylactic Effect of Probiotic Enterococcus lactis IW5 against Different Human Cancer Cells. Front Microbiol 6, 1317 (2015).

71. Kaeid Sharaf, L. & Shukla, G. Probiotics (Lactobacillus acidophilus and Lactobacillus rhamnosus GG) in Conjunction with Celecoxib (selective COX-2 inhibitor) Modulated DMH-Induced Early Experimental Colon Carcinogenesis. Nutr Cancer 70, 946–955 (2018).

72. Sharaf, L.K., Sharma, M., Chandel, D. & Shukla, G. Prophylactic intervention of probiotics (L.acidophilus, L.rhamnosus GG) and celecoxib modulate Bax-mediated apoptosis in 1,2-dimethylhydrazine-induced experimental colon carcinogenesis. BMC Cancer 18, 1111 (2018).

73. Li, J. et al. Probiotics modulated gut microbiota suppresses hepatocellular carcinoma growth in mice. Proc Natl Acad Sci U S A 113, E1306–1315 (2016).

74. Bajramagic, S. et al. Usage of Probiotics and its Clinical Significance at Surgically Treated Patients Sufferig from Colorectal Carcinoma. Med Arch 73, 316–320 (2019).

75. Azcarate-Peril, M.A., Sikes, M. & Bruno-Barcena, J.M. The intestinal microbiota, gastrointestinal environment and colorectal cancer: a putative role for probiotics in prevention of colorectal cancer? Am J Physiol Gastrointest Liver Physiol 301, G401–424 (2011).

76. Zhuo, Q. et al. Lysates of Lactobacillus acidophilus combined with CTLA-4-blocking antibodies enhance antitumor immunity in a mouse colon cancer model. Sci Rep 9, 20128 (2019).

77. Eeckhaut, V. et al. The Probiotic Butyricicoccus pullicaecorum Reduces Feed Conversion and Protects from Potentially Harmful Intestinal Microorganisms and Necrotic Enteritis in Broilers. Front Microbiol 7, 1416 (2016).

78. Everard, A. et al. Cross-talk between Akkermansia muciniphila and intestinal epithelium controls diet-induced obesity. Proc Natl Acad Sci U S A 110, 9066–9071 (2013).

79. Gopalakrishnan, V. et al. Gut microbiome modulates response to anti-PD-1 immunotherapy in melanoma patients. Science 359, 97–103 (2018).

80. Routy, B. et al. Gut microbiome influences efficacy of PD-1-based immunotherapy against epithelial tumors. Science 359, 91–97 (2018).

81. Zhai, R. et al. Strain-Specific Anti-inflammatory Properties of Two Akkermansia muciniphila Strains on Chronic Colitis in Mice. Front Cell Infect Microbiol 9, 239 (2019).

82. Paolillo, R., Romano Carratelli, C., Sorrentino, S., Mazzola, N. & Rizzo, A. Immunomodulatory effects of Lactobacillus plantarum on human colon cancer cells. Int Immunopharmacol 9, 1265–1271 (2009).

83. Kahouli, I., Malhotra, M., Westfall, S., Alaoui-Jamali, M.A. & Prakash, S. Design and validation of an orally administrated active L. fermentum-L. acidophilus probiotic formulation using colorectal cancer Apc (Min/+) mouse model. Appl Microbiol Biotechnol 101, 1999–2019 (2017).

84. Zhu, J. et al. Lactobacillus salivarius Ren prevent the early colorectal carcinogenesis in 1, 2-dimethylhydrazine-induced rat model. J Appl Microbiol 117, 208–216 (2014).

85. Chen, L. et al. NLRP12 attenuates colon inflammation by maintaining colonic microbial diversity and promoting protective commensal bacterial growth. Nat Immunol 18, 541–551 (2017).

86. Redman, M.G., Ward, E.J. & Phillips, R.S. The efficacy and safety of probiotics in people with cancer: a systematic review. Ann Oncol 25, 1919–1929 (2014).

87. Gibson, G.R. et al. Expert consensus document: The International Scientific Association for Probiotics and Prebiotics (ISAPP) consensus statement on the definition and scope of prebiotics. Nat Rev Gastroenterol Hepatol 14, 491–502 (2017).

88. van Loo, J., Coussement, P., de Leenheer, L., Hoebregs, H. & Smits, G. On the presence of inulin and oligofructose as natural ingredients in the western diet. Crit Rev Food Sci Nutr 35, 525–552 (1995).

89. Roberfroid, M.B., Van Loo, J.A. & Gibson, G.R. The bifidogenic nature of chicory inulin and its hydrolysis products. J Nutr 128, 11–19 (1998).

90. Campbell, J.M., Fahey, G.C., Jr. & Wolf, B.W. Selected indigestible oligosaccharides affect large bowel mass, cecal and fecal short-chain fatty acids, pH and microflora in rats. J Nutr 127, 130–136 (1997).

91. Fehlbaum, S. et al. In Vitro Fermentation of Selected Prebiotics and Their Effects on the Composition and Activity of the Adult Gut Microbiota. Int J Mol Sci 19 (2018).

92. Li, K. et al. Dietary inulin alleviates diverse stages of type 2 diabetes mellitus via anti-inflammation and modulating gut microbiota in db/db mice. Food Funct 10, 1915–1927 (2019).

93. Fuentes-Zaragoza, E. et al. Resistant starch as prebiotic: A review. Starch - Stärke 63, 406–415 (2011).

94. Sharma, A., Yadav, B.S. & Ritika Resistant Starch: Physiological Roles and Food Applications. Food Reviews International 24, 193–234 (2008).

95. Faraj, A., Vasanthan, T. & Hoover, R. The effect of extrusion cooking on resistant starch formation in waxy and regular barley flours. Food Research International 37, 517–525 (2004).

96. Zheng, D.W. et al. Prebiotics-Encapsulated Probiotic Spores Regulate Gut Microbiota and Suppress Colon Cancer. Adv Mater 32, e2004529 (2020).

97. Swanson, K.S. et al. The International Scientific Association for Probiotics and Prebiotics (ISAPP) consensus statement on the definition and scope of synbiotics. Nat Rev Gastroenterol Hepatol 17, 687–701 (2020).

98. Le Leu, R.K., Hu, Y., Brown, I.L., Woodman, R.J. & Young, G.P. Synbiotic intervention of Bifidobacterium lactis and resistant starch protects against colorectal cancer development in rats. Carcinogenesis 31, 246–251 (2010).

99. Ganesh, K. et al. Immunotherapy in colorectal cancer: rationale, challenges and potential. Nat Rev Gastroenterol Hepatol 16, 361–375 (2019).

100. Paulos, C.M. et al. Microbial translocation augments the function of adoptively transferred self/tumor-specific CD8+ T cells via TLR4 signaling. J Clin Invest 117, 2197–2204 (2007).

101. Iida, N. et al. Commensal bacteria control cancer response to therapy by modulating the tumor microenvironment. Science 342, 967–970 (2013).

102. Viaud, S. et al. The intestinal microbiota modulates the anticancer immune effects of cyclophosphamide. Science 342, 971–976 (2013).

103. Vetizou, M. et al. Anticancer immunotherapy by CTLA-4 blockade relies on the gut microbiota. Science 350, 1079–1084 (2015).

104. Sivan, A. et al. Commensal Bifidobacterium promotes antitumor immunity and facilitates anti-PD-L1 efficacy. Science 350, 1084–1089 (2015).

105. Gaines, S. et al. Western Diet Promotes Intestinal Colonization by Collagenolytic Microbes and Promotes Tumor Formation After Colorectal Surgery. Gastroenterology 158, 958–970 e952 (2020).

106. Chen, D.S. & Mellman, I. Elements of cancer immunity and the cancer– immune set point. Nature 541, 321–330 (2017).

107. Matson, V. et al. The commensal microbiome is associated with anti-PD-1 efficacy in metastatic melanoma patients. Science 359, 104–108 (2018).

108. Alberda, W.J. et al. The Importance of a Minimal Tumor-Free Resection Margin in Locally Recurrent Rectal Cancer. Dis Colon Rectum 58, 677–685 (2015).

109. Manfredi, S. et al. Incidence and patterns of recurrence after resection for cure of colonic cancer in a well defined population. Br J Surg 93, 1115–1122 (2006).

110. Naxerova, K. et al. Origins of lymphatic and distant metastases in human colorectal cancer. Science 357, 55–60 (2017).

111. Fermor, B., Umpleby, H.C., Lever, J.V., Symes, M.O. & Williamson, R.C. Proliferative and metastatic potential of exfoliated colorectal cancer cells. J Natl Cancer Inst 76, 347–349 (1986).

112. Shogan, B.D. et al. Collagen degradation and MMP9 activation by Enterococcus faecalis contribute to intestinal anastomotic leak. Sci Transl Med 7, 286ra268 (2015).

113. Mirnezami, A. et al. Increased local recurrence and reduced survival from colorectal cancer following anastomotic leak: systematic review and meta-analysis. Ann Surg 253, 890–899 (2011).

114. Moher, D., Liberati, A., Tetzlaff, J., Altman, D.G. & Group, P. Preferred reporting items for systematic reviews and meta-analyses: the PRISMA statement. PLoS Med 6, e1000097 (2009).

115. Shi, L. et al. Combining IL-2-based immunotherapy with commensal probiotics produces enhanced antitumor immune response and tumor clearance. J Immunother Cancer 8 (2020).

116. Arthur, J.C. et al. VSL#3 probiotic modifies mucosal microbial composition but does not reduce colitis-associated colorectal cancer. Sci Rep 3, 2868 (2013).

117. Chandel, D., Sharma, M., Chawla, V., Sachdeva, N. & Shukla, G. Isolation, characterization and identification of antigenotoxic and anticancerous indigenous probiotics and their prophylactic potential in experimental colon carcinogenesis. Sci Rep 9, 14769 (2019).

118. Song, H. et al. Pretreatment with probiotic Bifico ameliorates colitis-associated cancer in mice: Transcriptome and gut flora profiling. Cancer Sci 109, 666–677 (2018).

119. Xu, H. et al. Inhibitory Effects of Breast Milk-Derived Lactobacillus rhamnosus Probio-M9 on Colitis-Associated Carcinogenesis by Restoration of the Gut Microbiota in a Mouse Model. Nutrients 13 (2021).

120. Chang, S.C. et al. A gut butyrate-producing bacterium Butyricicoccus pullicaecorum regulates short-chain fatty acid transporter and receptor to reduce the progression of 1,2-dimethylhydrazine-associated colorectal cancer. Oncol Lett 20, 327 (2020).

121. Hradicka, P., Beal, J., Kassayova, M., Foey, A. & Demeckova, V. A Novel Lactic Acid Bacteria Mixture: Macrophage-Targeted Prophylactic Intervention in Colorectal Cancer Management. Microorganisms 8 (2020).

122. Silveira, D.S.C. et al. Lactobacillus bulgaricus inhibits colitis-associated cancer via a negative regulation of intestinal inflammation in azoxymethane/dextran sodium sulfate model. World J Gastroenterol 26, 6782–6794 (2020).

123. Yuan, L. et al. The influence of gut microbiota dysbiosis to the efficacy of 5-Fluorouracil treatment on colorectal cancer. Biomed Pharmacother 108, 184–193 (2018).

124. Chang, C.W. et al. Lactobacillus casei Variety rhamnosus Probiotic Preventively Attenuates 5-Fluorouracil/Oxaliplatin-Induced Intestinal Injury in a Syngeneic Colorectal Cancer Model. Front Microbiol 9, 983 (2018).

125. Bindels, L.B. et al. Increased gut permeability in cancer cachexia: mechanisms and clinical relevance. Oncotarget 9, 18224–18238 (2018).

126. Jacouton, E., Chain, F., Sokol, H., Langella, P. & Bermudez-Humaran, L.G. Probiotic Strain Lactobacillus casei BL23 Prevents Colitis-Associated Colorectal Cancer. Front Immunol 8, 1553 (2017).

127. Gao, C. et al. Gut Microbe-Mediated Suppression of Inflammation-Associated Colon Carcinogenesis by Luminal Histamine Production. Am J Pathol 187, 2323–2336 (2017).

128. Chung, E.J. et al. Combination of metformin and VSL#3 additively suppresses western-style diet induced colon cancer in mice. Eur J Pharmacol 794, 1–7 (2017).

129. da Silva Duarte, V. et al. Chemoprevention of DMH-Induced Early Colon Carcinogenesis in Male BALB/c Mice by Administration of Lactobacillus Paracasei DTA81. Microorganisms 8 (2020).

130. Mendes, M.C.S. et al. Microbiota modification by probiotic supplementation reduces colitis associated colon cancer in mice. World J Gastroenterol 24, 1995–2008 (2018).

131. Zhang, S.L. et al. Pectin supplement significantly enhanced the anti-PD-1 efficacy in tumor-bearing mice humanized with gut microbiota from patients with colorectal cancer. Theranostics 11, 4155–4170 (2021).

132. Li, Y. et al. Prebiotic-Induced Anti-tumor Immunity Attenuates Tumor Growth. Cell Rep 30, 1753–1766 e1756 (2020).

133. Zhu, L.Q. et al. Evodiamine inhibits high-fat diet-induced colitis-associated cancer in mice through regulating the gut microbiota. J Integr Med 19, 56–65 (2021).

134. Terasaki, M. et al. Alteration of fecal microbiota by fucoxanthin results in prevention of colorectal cancer in AOM/DSS-treated mice. Carcinogenesis (2020).

135. Luo, L. et al. The effect of menthol supplement diet on colitis-induced colon tumorigenesis and intestinal microbiota. Am J Transl Res 13, 38–56 (2021).

136. Liu, X., Wang, L., Jing, N., Jiang, G. & Liu, Z. Biostimulating Gut Microbiome with Bilberry Anthocyanin Combo to Enhance Anti-PD-L1 Efficiency against Murine Colon Cancer. Microorganisms 8 (2020).

137. Zhang, H., Hui, D., Li, Y., Xiong, G. & Fu, X. Canmei Formula Reduces Colitis-Associated Colorectal Carcinogenesis in Mice by Modulating the Composition of Gut Microbiota. Front Oncol 9, 1149 (2019).

138. Khan, I. et al. Mushroom polysaccharides and jiaogulan saponins exert cancer preventive effects by shaping the gut microbiota and microenvironment in Apc(Min/+) mice. Pharmacol Res 148, 104448 (2019).

139. Li, S., Fu, C., Zhao, Y. & He, J. Intervention with alpha-Ketoglutarate Ameliorates Colitis-Related Colorectal Carcinoma via Modulation of the Gut Microbiome. Biomed Res Int 2019, 8020785 (2019).

140. Fernandez, J. et al. A Galacto-Oligosaccharides Preparation Derived From Lactulose Protects Against Colorectal Cancer Development in an Animal Model. Front Microbiol 9, 2004 (2018).

141. Mudd, A.M. et al. Chemoprevention of Colorectal Cancer by Anthocyanidins and Mitigation of Metabolic Shifts Induced by Dysbiosis of the Gut Microbiome. Cancer Prev Res (Phila) 13, 41–52 (2020).

142. Wu, J.C. et al. Polymethoxyflavones prevent benzo[a]pyrene/dextran sodium sulfate-induced colorectal carcinogenesis through modulating xenobiotic metabolism and ameliorate autophagic defect in ICR mice. Int J Cancer 142, 1689–1701 (2018).

143. Wang, X. et al. Structural shift of gut microbiota during chemo-preventive effects of epigallocatechin gallate on colorectal carcinogenesis in mice. World J Gastroenterol 23, 8128–8139 (2017).

144. Chou, Y.C. et al. Boswellia serrata resin extract alleviates azoxymethane (AOM)/dextran sodium sulfate (DSS)-induced colon tumorigenesis. Mol Nutr Food Res 61 (2017).

145. Fukuda, M. et al. Prebiotic treatment reduced preneoplastic lesions through the downregulation of toll like receptor 4 in a chemo-induced carcinogenic model. J Clin Biochem Nutr 49, 57–61 (2011).

146. Oh, N.S., Lee, J.Y., Kim, Y.T., Kim, S.H. & Lee, J.H. Cancer-protective effect of a synbiotic combination between Lactobacillus gasseri 505 and a Cudrania tricuspidata leaf extract on colitis-associated colorectal cancer. Gut Microbes 12, 1785803 (2020).

147. Rudd, D.A. et al. Mapping insoluble indole metabolites in the gastrointestinal environment of a murine colorectal cancer model using desorption/ionisation on porous silicon imaging. Sci Rep 9, 12342 (2019).

148. Zhang, J. et al. A Functional Food Inhibits Azoxymethane/Dextran Sulfate Sodium-Induced Inflammatory Colorectal Cancer in Mice. Onco Targets Ther 14, 1465–1477 (2021).

149. Piazzi, G. et al. A Mediterranean Diet Mix Has Chemopreventive Effects in a Murine Model of Colorectal Cancer Modulating Apoptosis and the Gut Microbiota. Front Oncol 9, 140 (2019).

150. Li, F. et al. Modulation of colon cancer by nutmeg. J Proteome Res 14, 1937–1946 (2015).

151. Chen, L. et al. Chemoprevention of colorectal cancer by black raspberry anthocyanins involved the modulation of gut microbiota and SFRP2 demethylation. Carcinogenesis 39, 471–481 (2018).

152. Fernandez, J., Garcia, L., Monte, J., Villar, C.J. & Lombo, F. Functional Anthocyanin-Rich Sausages Diminish Colorectal Cancer in an Animal Model and Reduce Pro-Inflammatory Bacteria in the Intestinal Microbiota. Genes (Basel) 9 (2018).

153. Wang, C.Z. et al. Role of intestinal microbiome in American ginseng-mediated colon cancer prevention in high fat diet-fed AOM/DSS mice [corrected]. Clin Transl Oncol 20, 302–312 (2018).

154. Hu, Y. et al. Manipulation of the gut microbiota using resistant starch is associated with protection against colitis-associated colorectal cancer in rats. Carcinogenesis 37, 366–375 (2016).

155. Yu, C. et al. American ginseng significantly reduced the progression of high-fat-diet-enhanced colon carcinogenesis in Apc (Min/+) mice. J Ginseng Res 39, 230–237 (2015).

156. Gao, Z. et al. Probiotics modify human intestinal mucosa-associated microbiota in patients with colorectal cancer. Mol Med Rep 12, 6119–6127 (2015).

157. Zhang, J.W. et al. Preoperative probiotics decrease postoperative infectious complications of colorectal cancer. Am J Med Sci 343, 199–205 (2012).

158. Aisu, N. et al. Impact of perioperative probiotic treatment for surgical site infections in patients with colorectal cancer. Exp Ther Med 10, 966–972 (2015).

159. . Liu, Z., et al. Randomised clinical trial: the effects of perioperative probiotic treatment on barrier function and post-operative infectious complications in colorectal cancer surgery - a double-blind study. Aliment Pharmacol Ther 33, 50–63 (2011).

160. Gianotti, L. et al. A randomized double-blind trial on perioperative administration of probiotics in colorectal cancer patients. World J Gastroenterol 16, 167–175 (2010).

161. So, W.K.W. et al. Effects of a Rice Bran Dietary Intervention on the Composition of the Intestinal Microbiota of Adults with a High Risk of Colorectal Cancer: A Pilot Randomised-Controlled Trial. Nutrients 13 (2021).

162. Xie, X. et al. Effects of prebiotics on immunologic indicators and intestinal microbiota structure in perioperative colorectal cancer patients. Nutrition 61, 132–142 (2019).

163. Polakowski, C.B., Kato, M., Preti, V.B., Schieferdecker, M.E.M. & Ligocki Campos, A.C. Impact of the preoperative use of synbiotics in colorectal cancer patients: A prospective, randomized, double-blind, placebo-controlled study. Nutrition 58, 40–46 (2019).

164. Hibberd, A.A. et al. Intestinal microbiota is altered in patients with colon cancer and modified by probiotic intervention. BMJ Open Gastroenterol 4, e000145 (2017).

165. Worthley, D.L. et al. A human, double-blind, placebo-controlled, crossover trial of prebiotic, probiotic, and synbiotic supplementation: effects on luminal, inflammatory, epigenetic, and epithelial biomarkers of colorectal cancer. Am J Clin Nutr 90, 578–586 (2009).

166. Brown, D.G., Borresen, E.C., Brown, R.J. & Ryan, E.P. Heat-stabilised rice bran consumption by colorectal cancer survivors modulates stool metabolite profiles and metabolic networks: a randomised controlled trial. Br J Nutr 117, 1244–1256 (2017).

167. Nunez-Sanchez, M.A. et al. Gene expression changes in colon tissues from colorectal cancer patients following the intake of an ellagitannin-containing pomegranate extract: a randomized clinical trial. J Nutr Biochem 42, 126–133 (2017).

168. Nunez-Sanchez, M.A. et al. MicroRNAs expression in normal and malignant colon tissues as biomarkers of colorectal cancer and in response to pomegranate extracts consumption: Critical issues to discern between modulatory effects and potential artefacts. Mol Nutr Food Res 59, 1973–1986 (2015).

169. Chen, C.C. et al. Oral inoculation of probiotics Lactobacillus acidophilus NCFM suppresses tumour growth both in segmental orthotopic colon cancer and extra-intestinal tissue. Br J Nutr 107, 1623–1634 (2012).

170. Bellville, J.W., Swanson, G.D., Miyake, T. & Aqleh, K.A. Respiratory stimulation observed following ethanol ingestion. West J Med 124, 423–425 (1976).

171. Shinohara, K., Ohashi, Y., Kawasumi, K., Terada, A. & Fujisawa, T. Effect of apple intake on fecal microbiota and metabolites in humans. Anaerobe 16, 510–515 (2010).

172. Schwartz, S.E. et al. Sustained pectin ingestion: effect on gastric emptying and glucose tolerance in non-insulin-dependent diabetic patients. Am J Clin Nutr 48, 1413–1417 (1988).

173. Annunziata, G. et al. Effect of Grape Pomace Polyphenols With or Without Pectin on TMAO Serum Levels Assessed by LC/MS-Based Assay: A Preliminary Clinical Study on Overweight/Obese Subjects. Front Pharmacol 10, 575 (2019).

174. Glinsky, V.V. & Raz, A. Modified citrus pectin anti-metastatic properties: one bullet, multiple targets. Carbohydr Res 344, 1788–1791 (2009).

175. Warburg, O. On the origin of cancer cells. Science 123, 309–314 (1956).

176. Roediger, W.E. Role of anaerobic bacteria in the metabolic welfare of the colonic mucosa in man. Gut 21, 793–798 (1980).

177. Khoo, H.E., Azlan, A., Tang, S.T. & Lim, S.M. Anthocyanidins and anthocyanins: colored pigments as food, pharmaceutical ingredients, and the potential health benefits. Food Nutr Res 61, 1361779 (2017).

178. Reglero, C. & Reglero, G. Precision Nutrition and Cancer Relapse Prevention: A Systematic Literature Review. Nutrients 11 (2019).

179. Toledo, E. et al. Mediterranean Diet and Invasive Breast Cancer Risk Among Women at High Cardiovascular Risk in the PREDIMED Trial: A Randomized Clinical Trial. JAMA Intern Med 175, 1752–1760 (2015).

180. Bamia, C. et al. Mediterranean diet and colorectal cancer risk: results from a European cohort. Eur J Epidemiol 28, 317–328 (2013).

181. (2018).

182. Henderson, A.J. et al. Chemopreventive properties of dietary rice bran: current status and future prospects. Adv Nutr 3, 643–653 (2012).

